# A digital biomarker of white-matter routing from diffusion MRI as an early biomarker of Alzheimer’s disease

**DOI:** 10.64898/2026.10.21.26366165

**Authors:** Merit Ghodrat, Johannes CM Schlachetzki, the Alzheimer’s Disease Neuroimaging Initiative

## Abstract

Alzheimer’s disease is diagnosed after a decade-long phase in which pathology accumulates; current markers show that pathology is present, but not how far it has advanced. Because white-matter structure is altered early, we computed two graph-theoretic indices of periventricular white-matter routing from diffusion MRI in 1,706 OASIS-3 sessions: the Route Concordance Index (RCI), a population-calibrated path-concordance score over 316 routes, and a global conductance from the connectome’s graph Laplacian. RCI was reduced before clinical conversion and declined monotonically across a five-stage trajectory (cross-validated AUC = 0.817 for stable-CN vs. pre-AD; comparable to hippocampal volume and exceeding entorhinal thickness, global conductance, and DTI-ALPS), with degradation concentrated in fronto-striatal circuits. A classifier combining these routing features with demographic, structural-MRI and cognitive covariates detected conversion to MCI or AD up to four years before diagnosis (AUC = 0.831; 0.918 within one year; modest positive predictive value). Findings replicated in ADNI (*n* = 609; conductance AUC = 0.805; RCI = 0.797 under frozen external validation) and tracked CSF amyloid-*β*_42_ in the expected direction. Computed from a standard diffusion MRI sequence, RCI is comparable to established structural and demographic biomarkers on preclinical discrimination while, unlike them, declining cleanly and monotonically across the disease trajectory, supporting prospective validation for risk stratification.

## Introduction

Alzheimer’s disease (AD) is preceded by a decade-long preclinical phase during which amyloid-*β* and hyperphosphorylated tau accumulate before measurable cognitive decline ^1, 2^. Current markers, such as CSF amyloid-*β*42, amyloid-PET, and plasma pTau217 ^2^, establish that pathology is present but not how far it has advanced. Diffusion-MRI measures the displacement of water and is sensitive to medial-temporal and posterior microstructural changes that track Alzheimer’s neuropathology and precede dementia ^3, 4^. Detecting this phase non-invasively, at scale, and without specialized fluid or PET biomarkers remains an unmet need. Digital biomarkers, objective measures computed algorithmically from routinely collected data ^5^, are well suited to this need.

The brain lacks conventional lymphatic vessels and clears interstitial solutes, including amyloid and tau species, by a combination of diffusion and fluid movement along perivascular spaces and meningeal and cranial exit routes ^6, 7^. Impairment of brain clearance has been implicated in AD ^8, 9^ through the relative contributions of convective flow and diffusion. Recent tracing of endogenous neuronal proteins indicates that clearance is regionally compartmentalised, with different brain regions draining by different routes ^10^. Whatever the balance of processes, solute movement occurs along perivascular and white-matter routes whose structural integrity is measurable with diffusion MRI.

The most widely used non-invasive proxy is the diffusion tensor image analysis along the perivascular space (DTI-ALPS) index, which estimates perivascular water mobility from diffusivity along three orthogonal axes in a small region of interest near the lateral ventricle ^11^. It summarizes diffusion at a single periventricular location; here we instead take a network-level view of white-matter routing.

We hypothesized that brains of cognitively normal individuals traverse reproducible, population-typical white matter routes from periventricular CSF sources to cortical and subcortical targets, and that this routing signature is progressively disrupted across the AD trajectory. To test this, we built a new digital biomarker of periventricular white-matter routing integrity: the Route Concordance Index (RCI), a population-calibrated path-concordance score, paired with a companion graph-Laplacian global-conductance metric derived from clinical DTI through the fully automated pipeline outlined in Figure 1. Both are instances of a more general construction: a trainable, graph-theoretic index that scores how closely an individual’s structural-routing signature matches a reference population’s.

**Figure 1:**
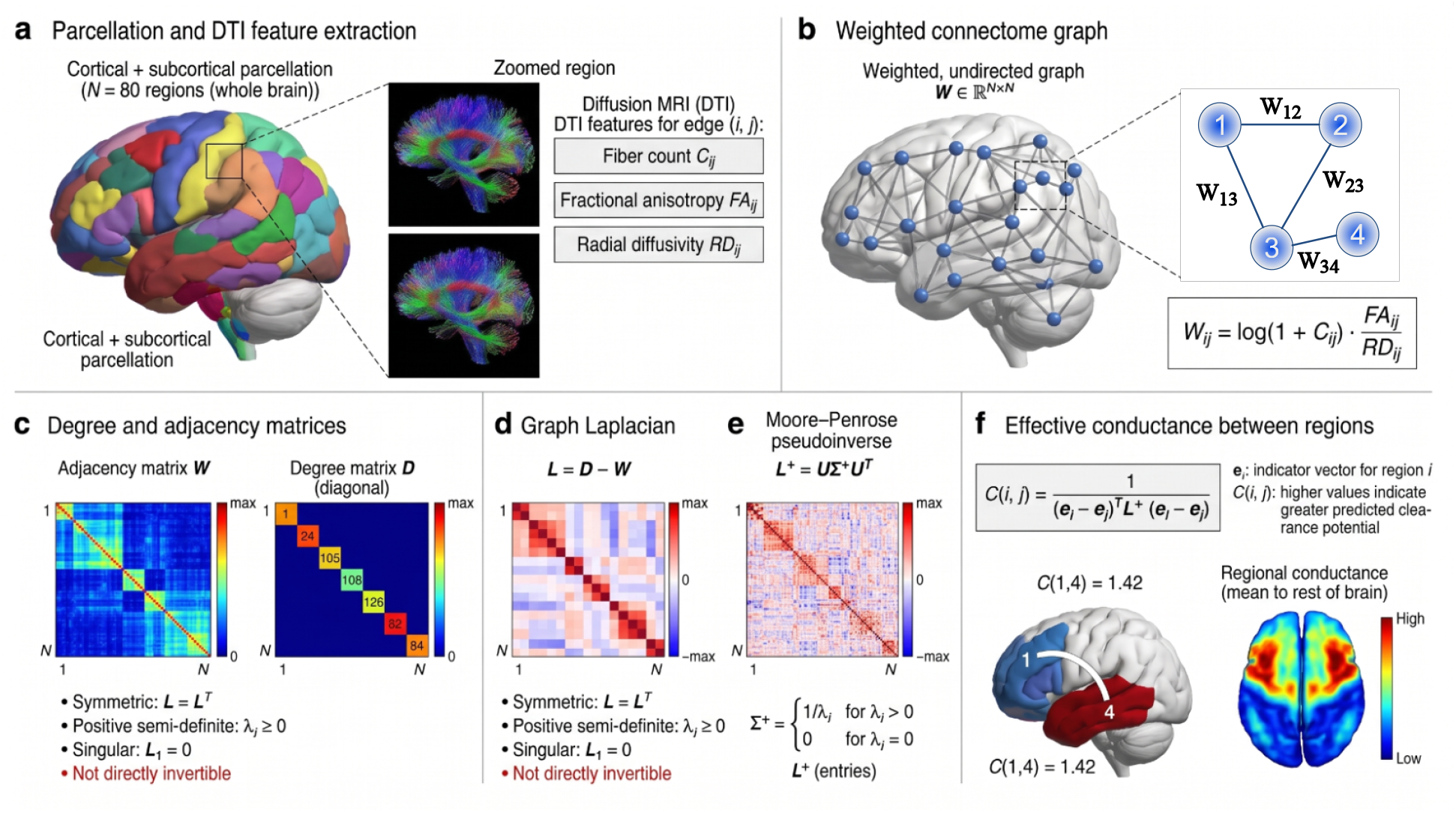
Effective-conductance pipeline from standard DTI. Each of the 80×80 region-pair DTI measurements (streamline count, fractional anisotropy, radial diffusivity) is compressed into a single edge weight, assembled into a weighted connectome graph, converted to a graph Laplacian, and inverted via its Moore–Penrose pseudoinverse to obtain the Kirchhoff effective conductance between any source and target region, a quantity that integrates contributions from all parallel white-matter pathways simultaneously, not just the single strongest route. Four periventricular regions (bilateral thalamus and caudate), candidate sites of perivascular CSF entry ^8^, were paired with each of the remaining 79 Desikan–Killiany–Tourville/FreeSurfer atlas regions ^25, 26, 27^, yielding 316 anatomically grounded source→target routes per session, reconstructed from whole-brain deterministic tractography ^28^. The same 316 routes are the input to the route-enumeration stage in Supplementary Fig. S1.

Here we show that RCI separates diagnostic stages more cleanly than established markers. RCI was the only index to decline monotonically across the full five-stage cognitive trajectory, and it diverged from the cognitively normal reference earlier than hippocampal volume. Its degradation was anatomically selective, concentrated in fronto-striatal routes while the canonical hippocampal-fornix backbone remained comparatively preserved, rather than tracking diffuse atrophy. The effect replicated in an independent, 53-site cohort and moved in the expected direction with CSF amyloid-*β*_42_. Together these results establish RCI as a monotonic, anatomically specific, and externally replicated digital marker of early Alzheimer’s-related structural change.

## Results

### RCI is the only structural-routing feature that declines cleanly monotonically across the Alzheimer’s trajectory

Here we asked, whether individuals who later convert to MCI or AD already show distinct white-matter routing before clinical conversion, compared with cognitively normal (CN) individuals who do not convert. To address this question, we computed two indices of periventricular white-matter routing: RCI and global conductance, derived from 1,706 OASIS-3 sessions and 609 ADNI sessions used for discovery and validation, respectively. Whole-brain deterministic tractography over an 80-region atlas yielded the structural connectome per session. Four periventricular regions, i.e., bilateral thalamus and caudate, adjacent to candidate sites of perivascular CSF entry were paired with each of the remaining 79, giving 316 source-to-target routes (Supplementary Fig. S1). RCI scores how closely the routes an individual uses match those preferentially used by the CN reference population, weighted by each route’s structural quality. Global conductance treats the same connectome as a resistor network and measures, for each of the same 316 pairs, the total flow that all parallel pathways together would support. The two use identical routes but are derived independently, so their agreement is informative rather than automatic. We assigned sessions to five mutually exclusive groups from longitudinal Clinical Dementia Rating (CDR) trajectories: (1) stable CN, CDR=0 with no conversion between imaging visits; (2) pre-clinical MCI, CDR=0 at earlier imaging visits, converting to MCI at a later imaging visit; (3) pre-clinical AD, CDR ≤ 0.5 at earlier imaging visits, converting to AD (CDR ≥ 1) at a later visit; (4) post-diagnostic MCI and (5) post-diagnostic AD with CDR = 0.5 and CDR ≥ 1.0, respectively, imaged after a documented conversion to MCI or dementia without prior imaging visits as CN. All three structural-routing features separated stable CN from every disease stage (Figure 2; Kruskal–Wallis *p* < 0.001 for every feature; pairwise stable-CN-vs-group contrasts significant at *p* < 0.001 for conductance and RCI at every disease stage), but they differ in whether they behave as staging metrics. Radial diffusivity (Fig. 2A) and global conductance (Fig. 2B) both show a reversal: pre-clinical AD sessions are numerically worse (higher RD, lower conductance) than both post-diagnostic groups, so neither feature is monotonic across the full five-stage ordering. RCI (Fig. 2C) is the only one of the three routing features that declines cleanly and monotonically at every stage transition; the remaining anatomical and predictive analyses below use RCI as the primary metric. Taken together, RCI was already lower at imaging visits acquired before a later conversion, and was the only measure to decline at every disease stage.

**Figure 2:**
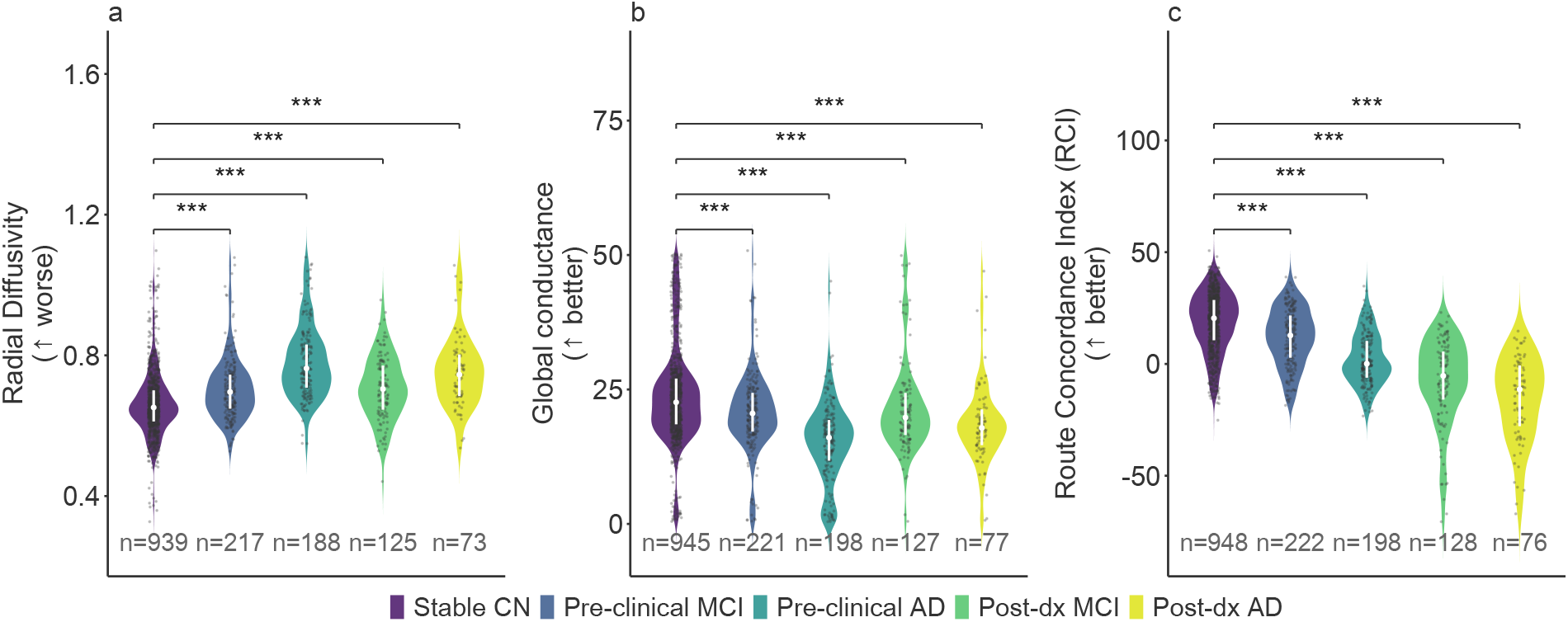
Structural white-matter routing features across five diagnostic trajectory groups in OASIS-3. Stable CN, pre-clinical MCI, pre-clinical AD, post-diagnostic MCI, and post-diagnostic AD (group *n* shown per panel), after independent Tukey 3×IQR outlier pruning within each panel (panel-specific *N* and outliers removed reported in each subtitle; pruning is not shared across features because each has a different missingness/outlier pattern). (A) Radial diffusivity and (B) global conductance both separate stable CN from every other stage, but neither is monotonic across all five stages: pre-clinical AD is numerically worse than both post-diagnostic stages for these two features. (C) RCI is the only feature that declines cleanly and monotonically across all five stages. Asterisks denote Bonferroni-corrected Mann–Whitney *U* comparisons against the stable-CN group (*** *p* < 0.001, ** *p* < 0.01, * *p* < 0.05); inset boxes report omnibus Kruskal– Wallis significance.

### RCI is comparable to established structural and demographic biomarkers while staging the disease monotonically

We benchmarked RCI against 11 established markers (Figure 3). For stable CN versus pre-clinical AD, residualized RCI reached a cross-validated AUC of 0.817 [95% CI 0.788–0.847] (Fig. 3A; concordance weights fit within each fold, see Methods), statistically indistinguishable from hippocampal volume (0.804; paired DeLong *p* = 0.49) and radial diffusivity (0.822; *p* = 0.81), and significantly higher than entorhinal thickness (0.687), global conductance (0.713), and DTI-ALPS (0.518, at chance; all *p* < 10*^−^*^5^; Supplementary Table S2). Unlike radial diffusivity, however, RCI was the only leading marker to order all five disease stages monotonically (Figure 2). Once disease was clinically manifest, atrophy markers were at least as discriminative (pre-versus post-diagnostic AUC 0.592 for RCI versus 0.688 for hippocampal volume; Fig. 3B), placing RCI’s contribution in the preclinical window. The result held within each acquisition protocol (single-shell DTI 0.815, DSI 0.838, multishell 0.721). Taken together, the pre-clinical contrast RCI matched the best established structural biomarkers and demographic risk factors while being the only one to stage the disease monotonically.

**Figure 3:**
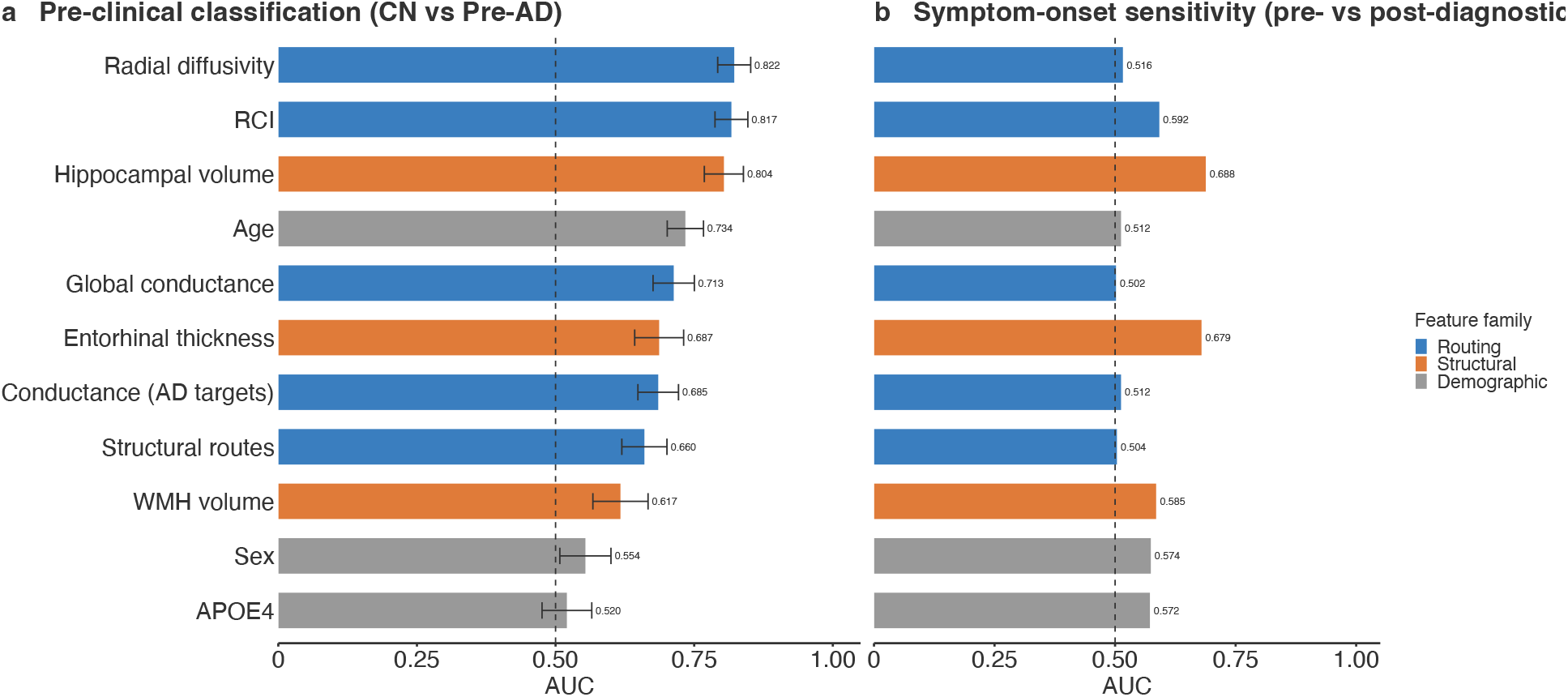
RCI matches the leading biomarkers on pre-clinical classification but not on symptom-onset sensitivity. All imaging and structural biomarkers are age/sex/APOE4-residualized (CN-only normative OLS model); the demographic and genetic markers (age, sex, APOE4) are shown unadjusted, since a variable cannot be residualized against itself. Bars are AUC (Mann–Whitney *U* formulation) with DeLong 95% CI, colored by category: structural-routing metrics from this study (blue), established AD structural biomarkers (orange), and demographic/genetic covariates (gray). AUCs are oriented to the discriminative direction, max(AUC, 1− AUC); markers that increase rather than decrease with disease (e.g. radial diffusivity, white-matter hyperintensity volume) are flipped accordingly, so every bar lies at or above 0.5. (A) Pre-clinical classification (stable CN vs. pre-clinical AD): radial diffusivity (0.822), RCI (0.817), and hippocampal volume (0.804) lead, followed by age (0.781), global conductance (0.713), entorhinal cortical thickness (0.687), conductance restricted to AD-target regions (0.685), the count of structural routes present (0.660), white-matter hyperintensity volume (0.617), APOE4 (0.588), and sex (0.551). RCI and radial diffusivity do not differ (paired DeLong *p* = 0.81); as noted in the text, RCI’s advantage over that marker is one of monotonic staging rather than of discrimination. (B) Symptom-onset sensitivity (pre-vs. post-diagnostic AUC, averaged across the MCI and AD tracks). Under cross-validated RCI weights this within-disease discrimination is modest (AUC= 0.592), below hippocampal volume (0.688) and entorhinal thickness (0.679). RCI’s advantage is thus specific to the preclinical stable-CN-vs-pre-AD contrast (panel A), where neither group defines the concordance weights, rather than to staging established disease, consistent with morphometric atrophy becoming more discriminative once the disease is symptomatic.

### Disease-specific degradation concentrates in clearance-relevant anatomical circuits

If routing degradation were a non-specific consequence of global neurodegeneration, the spatial pattern of route loss would be expected to track overall white-matter burden rather than a specific anatomical distribution. Instead, route-level analysis (Figure 4) showed that the hippocampal-fornix system (the canonical periventricular-to-medial-temporal pathway and the earliest site of neurofibrillary tangle accumulation in Braak staging ^12^) remains the single most preserved part of the network even at the post-diagnostic AD stage (prevalence 86–99% → 73–93%, a 6–13 percentage-point decline; Fig. 4A), while fronto-striatal routes, which are not classically implicated as the earliest AD circuits, degrade far more (a 33–37 percentage-point decline in route prevalence and 32–52% decline in conditional conductance; Fig. 4B). This anatomical pattern, broad frontostriatal vulnerability with relative preservation of the canonical hippocampal-fornix backbone, is consistent with a distributed network-level process. Collectively, route loss was anatomically structured rather than diffuse, concentrating in fronto-striatal connections while relatively sparing the hippocampal-fornix pathway.

**Figure 4:**
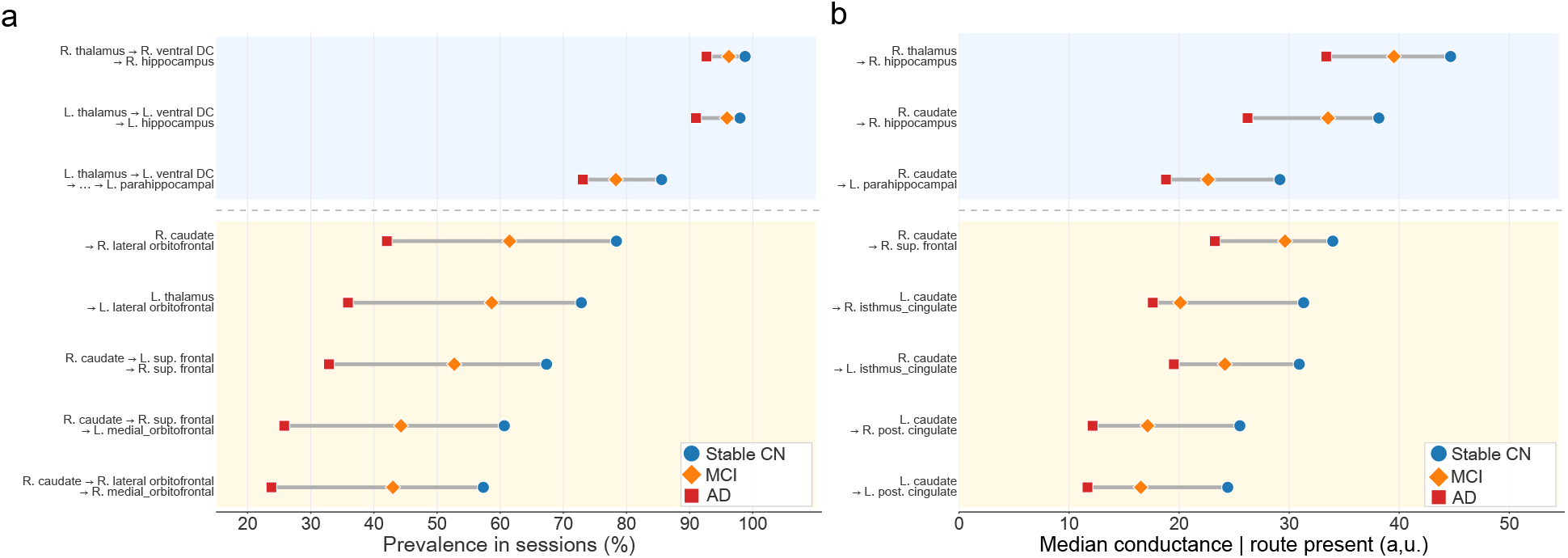
Anatomical specificity of white-matter route degradation. (A) Prevalence of the canonical (most structurally favored) route for each source→target pair, by diagnostic group. Hippocampal/fornix routes (thalamus/ventral diencephalon → hippocampus and parahippocampal gyrus) are used by 86–99% of stable-CN sessions and decline only modestly toward AD (6–13 percentage points), remaining the most robust routes in the network even at the AD stage. Fronto-striatal routes (caudate/thalamus → orbitofrontal and superior frontal cortex) are comparatively fragile, declining 33–37 percentage points from CN to AD. (B) Median conductance conditional on route presence, ranked by the magnitude of CN→AD degradation; hippocampal-fornix routes generally show a smaller percentage decline (25–36%) than fronto-striatal routes (32–52%). For transparency: route prevalence and conductance are computed from raw streamline tractography without correction for tractography’s known tendency to overcount certain connection types; because this bias applies uniformly across the diagnostic groups, the between-group contrasts reported here are not affected by it.

### White-matter routing decline precedes clinical diagnosis and intensifies with disease progression

Because preclinical AD is defined operationally by biomarker evidence of pathology in the absence of overt cognitive impairment ^1, 2^, the clinical utility of a candidate biomarker depends not only on its overall group separation but on how early it diverges from cognitively normal. Figure 5a places each biomarker on two axes, overall group separation (Kruskal–Wallis *ε*^2^) and monotonic-trend strength (Jonckheere–Terpstra *z*); RCI has the strongest monotonic trend (*z* = 16.9) and the second-largest separation (*ε*^2^ = 0.193, after hippocampal volume at 0.204), marking it as both discriminative and cleanly monotonic. The staging curves in Figure 5b then show that RCI diverges from the CN reference earliest among the top six biomarkers, with a visibly steeper initial slope at the pre-clinical MCI and pre-clinical AD stages than hippocampal volume or entorhinal thickness, both of which show their primary departure later, at the post-diagnostic stages. This pattern is consistent with the hypothesis that white-matter routing disruption is an early, rather than terminal, feature of the AD trajectory. Four of the other five biomarkers in Figure 5b (hippocampal volume, global conductance, conductance to AD-target regions, and structural routes present) dip at the pre-clinical AD stage and partially recover at the post-diagnostic MCI stage before declining further at post-diagnostic AD; RCI is again the only biomarker with no such reversal. To summarize, RCI changed at the earliest disease stage and without reversal, whereas hippocampal volume and entorhinal thickness separated from the reference only after diagnosis.

**Figure 5:**
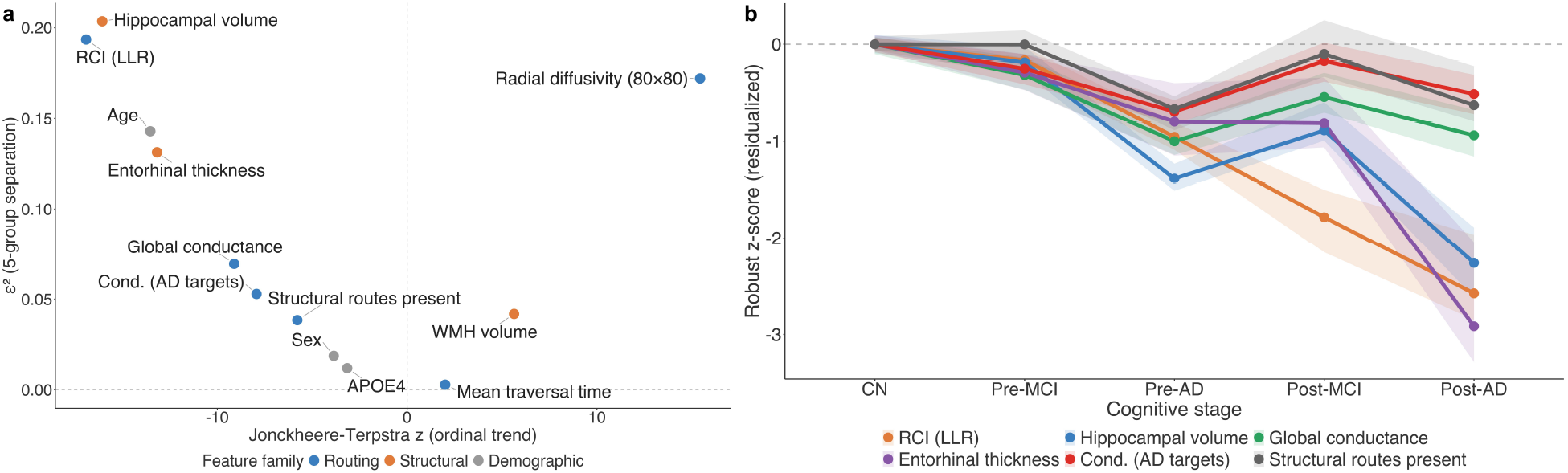
Staging trajectory of the top biomarkers. (a) A monotonicity-versus-separation comparison. The horizontal axis is the Kruskal–Wallis effect size (*ε*^2^), which measures how strongly a biomarker separates the five groups overall and is therefore relevant to case detection and classification; the vertical axis is the Jonckheere–Terpstra trend statistic, which measures whether a biomarker changes monotonically across the ordered disease stages and is therefore relevant to disease staging and tracking progression. RCI scores high on both, indicating it is useful both as a discriminative classifier and as a monotonic staging marker. Shown alongside (b) age/sex/APOE4-adjusted staging curves (median and bootstrap 95% CI, robust *z*-standardized relative to the CN reference) for the top six biomarkers across the five ordinal disease stages. RCI shows the earliest and steepest divergence from the CN reference, already below CN at the pre-clinical MCI stage in the adjusted curves (a difference that does not survive age/sex propensity matching; Supplementary Table S1) and before hippocampal volume, entorhinal thickness, or global conductance show comparable separation, consistent with white-matter routing impairment preceding measurable structural atrophy.

### A routing-based machine learning model detects conversion up to four years before diagnosis

Table 1 summarizes lead-time-stratified detection performance (Fig. 6A, B). Discrimination was strongest closest to diagnosis and degraded gracefully, rather than collapsing, with increasing lead time, a pattern consistent with biomarker evidence accumulating progressively rather than appearing abruptly at conversion. Positive predictive value is modest at any fixed operating point in a low-prevalence setting: at a high-precision (PPV≥80%) threshold, sensitivity falls to 0.41–0.56. Because positive predictive value scales with prevalence, its value in an unselected screening population would be lower than in this enriched cross-validation cohort. The limitation is not primarily one of evidence strength but of operating point and base rate, and it means the model as presented is better suited to enriching a higher-risk sub-population (e.g. a memory-clinic referral) for further workup than to standalone, unenriched population screening. An ordinal extension of the model, which predicts disease stage rather than a binary conversion label using a Frank–Hall decomposition ^13^, achieved a pre-clinical discrimination AUC of 0.809 and a Spearman rank correlation of *ρ* = 0.536 (*p* = 1.0 × 10*^−^*^26^) between predicted ordinal stage and true class, with 85.3% adjacent-class accuracy. A feature-ablation analysis confirmed the model is not a proxy for age or genotype: a classifier trained on the structural-routing features alone reached ROC-AUC 0.813 [95% CI 0.787, 0.839], and adding age, sex, and APOE4 did not improve it (0.812 [0.787, 0.838]); demographic risk factors carry essentially no discriminative information beyond the structural-routing features.

**Figure 6:**
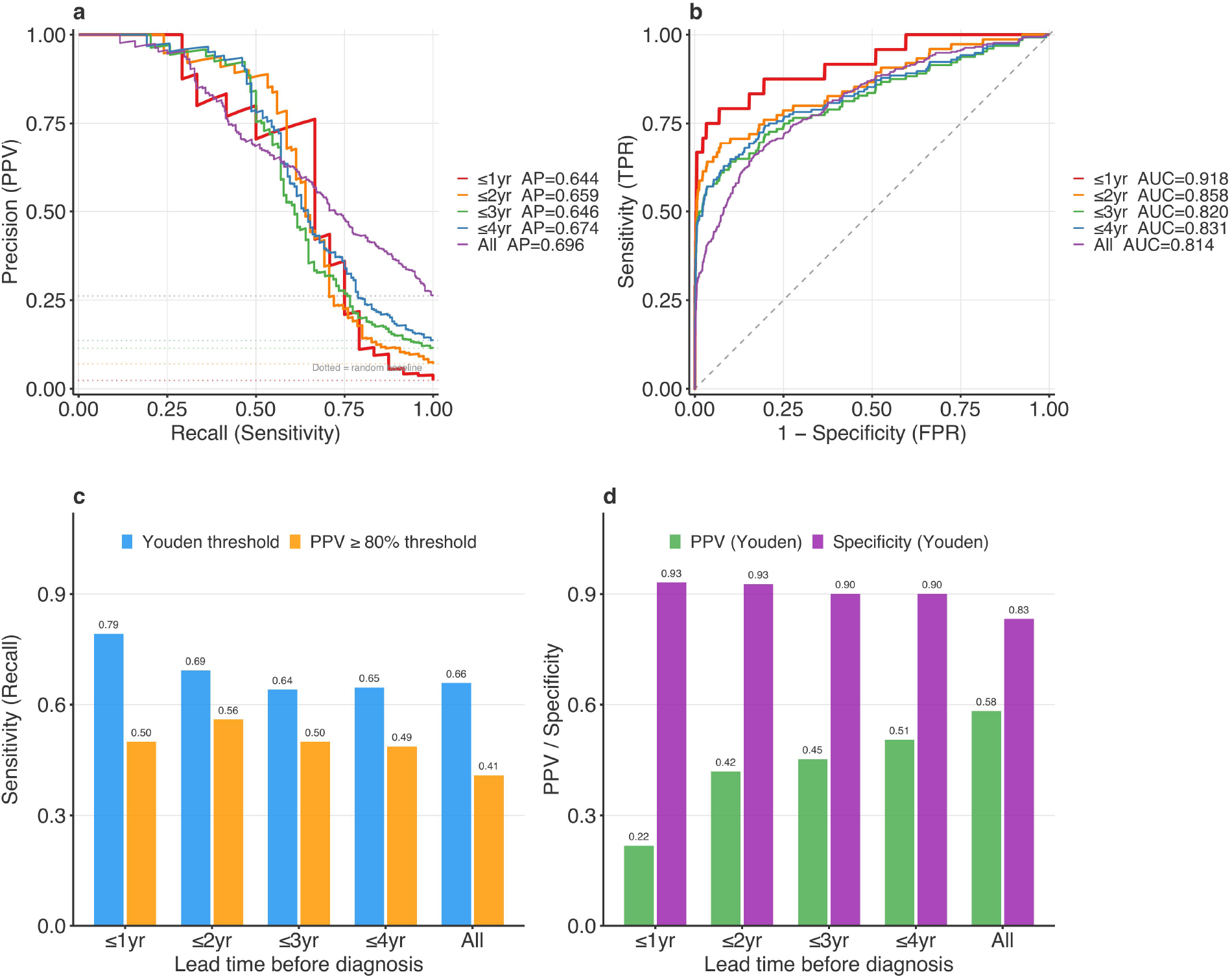
Preclinical detection performance by lead time before clinical diagnosis. Evidence of analytical utility: a histogram-based gradient-boosted classifier (a standard, off-the-shelf algorithm, implemented via the scikit-learn library ^35^) trained on our structural-routing feature set (seven families of 316 edge-level *z*-scores: conductance, streamline count, FA, RD, per-edge RCI, top-path score, and route entropy; 80 traversal-time features; and demographic, structural-MRI, and cognitive covariates; 2,306 columns total) under repeated subject-grouped cross-validation, with within-fold SMOTE class rebalancing ^36^ and RCI log-likelihood weights re-estimated separately in every training fold to prevent leakage. (A) Precision–recall and (B) ROC curves stratified by the number of years between the MRI session and eventual clinical diagnosis. (C) Discrimination (AUC, PR-AUC) as a function of lead time. (D–F) Sensitivity, specificity, and PPV at the Youden-optimal and PPV≥80% operating points. AUC was highest closest to diagnosis (0.918 at ≤1 year, *n* = 24 converters) and remained well above chance across the full four-year preclinical window (0.814 across all 352 pre-clinical sessions; 0.831 restricted to conversions within 4 years, *n* = 156), without PET or fluid biomarkers.

**Table 1:** Preclinical Alzheimer’s detection performance by lead time before clinical diagnosis (OASIS-3, pooled out-of-fold predictions from leakage-corrected probabilities, *n*_stable CN_ = 991). The 352 pre-clinical sessions in the “All pre-clinical” row are the pre-MCI and pre-AD sessions (of 435 total) that had a documented conversion date and thus a definable lead time; the remaining 83 lacked a datable conversion and were not used as positives. Sensitivity and specificity are prevalence-independent operating characteristics at the Youden-optimal threshold. Positive predictive value is prevalence-dependent and is therefore not tabulated at the (non-representative) cross-validation prevalence; at the all-pre-clinical operating point (sensitivity 0.66, specificity 0.83) it corresponds to approximately 17% at a 5% screening prevalence and 63% at a 30% memory-clinic prevalence.

| Lead window | $N$ converters | AUC | PR-AUC | Sensitivity (Youden) | Specificity (Youden) |
| --- | --- | --- | --- | --- | --- |
| $\leq 1$ year | 24 | 0.918 | 0.644 | 0.792 | 0.931 |
| $\leq 2$ years | 75 | 0.858 | 0.659 | 0.693 | 0.927 |
| $\leq 3$ years | 128 | 0.820 | 0.646 | 0.641 | 0.900 |
| $\leq 4$ years | 156 | 0.831 | 0.674 | 0.647 | 0.900 |
| All pre-clinical | 352 | 0.814 | 0.696 | 0.659 | 0.832 |

### External replication in ADNI and association with CSF amyloid-*β*_42_

The main results replicated in an independent ADNI cohort (Figure 7, ComBat-harmonized, n = 609), acquired on different scanners at different sites with no overlapping participants. Global conductance discriminated CN from AD at AUC= 0.805 [95% CI 0.710–0.900], and RCI, with its weights re-estimated within ADNI, at AUC= 0.909 [0.867–0.950], both in the same CN > MCI > AD direction seen in OASIS-3. The same effects held without harmonization in the paired 2mm sensitivity stratum (conductance 0.815 [0.694–0.935]; RCI 0.898 [0.843–0.953]; all contrasts and strata in Supplementary Table S3). Because global conductance is untrained, its ADNI performance is a direct external validation; the RCI weights above, however, were re-estimated within ADNI, so they show that the method reproduces rather than that the OASIS model itself transfers. Applying the OASIS-trained RCI weights frozen to ADNI, a stricter external-validation test, the OASIS model still discriminated CN from AD (AUC= 0.797 [95% CI 0.725, 0.868]) and CN from MCI (AUC= 0.633 [0.585, 0.681]), confirming genuine cross-cohort transfer with the attenuation expected when weights are not re-fit. Testing convergence with molecular markers (Supplementary Fig. S2), the strongest and only metric-consistent signal was CSF amyloid-*β*_42_, which correlated with both RCI (*ρ* = +0.499, *p* = 0.003, *n* = 34, paired 2mm stratum) and conductance (*ρ* = +0.542, p = 0.001) in the expected direction, since lower CSF amyloid-β_42_ reflects more amyloid retained in brain parenchyma. Plasma pTau-217 correlated with RCI in the predicted direction in the paired 2mm stratum (*ρ* = −0.235, *p* = 0.035, *n* = 81), but not in the full harmonized cohort (*ρ* = −0.156, *p* = 0.074, *n* = 132), where conductance instead reached significance (*ρ* = −0.189, *p* = 0.030). Continuous amyloid-PET and tau-PET burden were not significantly associated with either metric; a raw inverse RCI-to-amyloid-PET association in OASIS-3 (*ρ* = −0.149, *p* < 0.001, *n* = 1,313 sessions from 726 participants, on the Centiloid scale ^14^) did not survive age/sex/APOE4 adjustment (partial *ρ* = +0.02, *p* = 0.37), and the ADNI amyloid-PET measure is a pooled multitracer SUVR rather than a tracer-harmonized Centiloid value, so at *n* = 54 this null should be read cautiously. CSF pTau was not associated with either metric in the paired 2mm stratum (*n* = 34); RCI showed a positive correlation with CSF pTau in the full cohort (*ρ* = +0.365, *p* = 0.005, *n* = 57), but this is opposite to the direction the clearance hypothesis predicts and is uncorrected for the 28 Tier-B correlations tested (Supplementary Table S4). As specificity checks, APOE4 copy number showed no dose-response with either index (*ρ* < 0.08, *p* > 0.2, *n* ≈ 284–418), while allele dose at rs12454744, upstream of AQP4 ^15^, showed a nominal positive relationship with RCI (*ρ* = 0.196, *p* = 0.020, *n* = 142) but not conductance; this did not survive Bonferroni correction and is hypothesis-generating. To summarize, RCI and global conductance validated the CN-MCI-AD gradient in the independent ADNI cohort and tracked CSF amyloid-*β*_42_.

**Figure 7:**
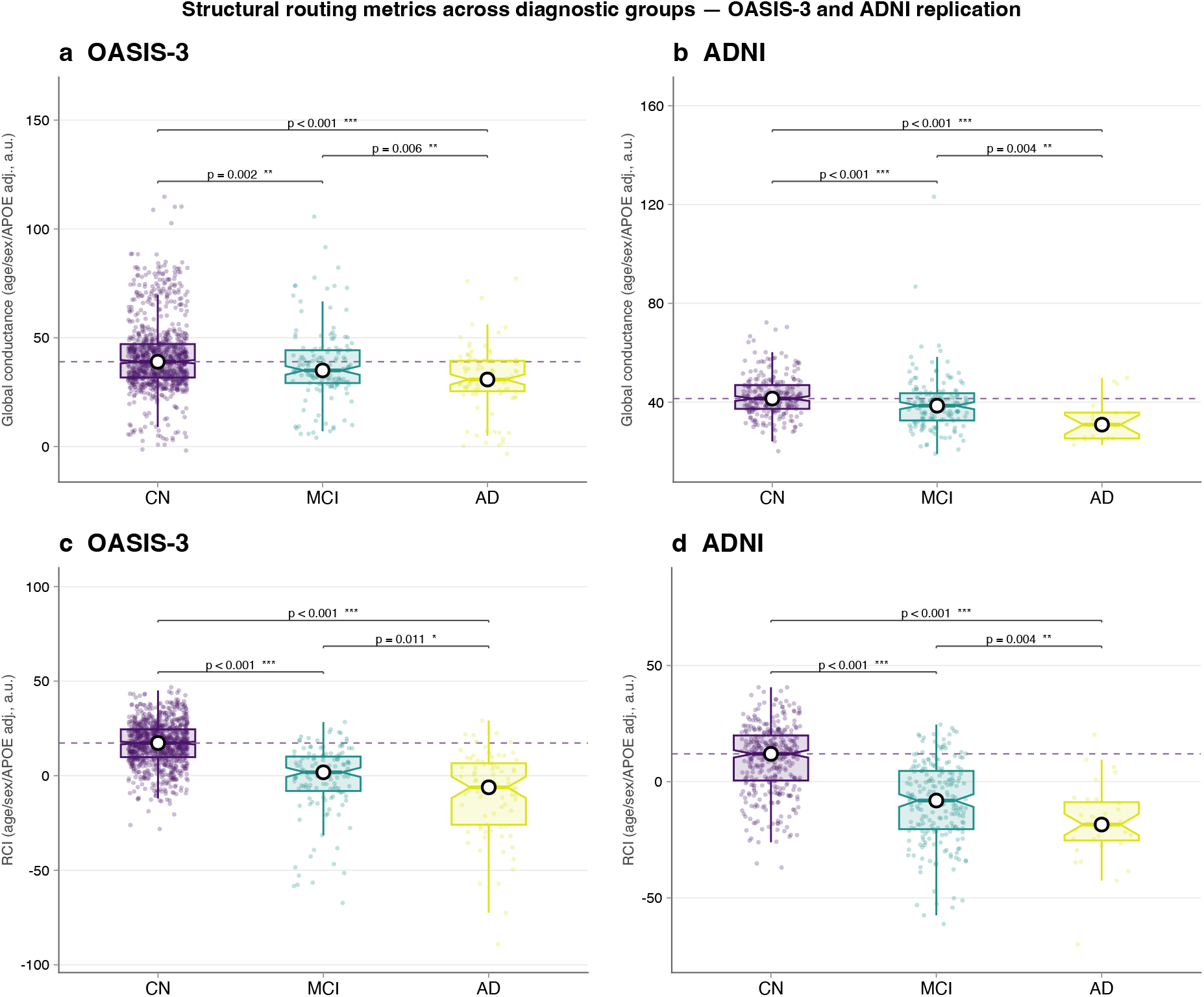
External replication across two independent cohorts. Age/sex/APOE4-adjusted (A, B) global conductance and (C, D) RCI across CN/MCI/AD groups in OASIS-3 (A, C) and ADNI (B, D). OASIS-3 groups are stable-CN, post-diagnostic MCI, and post-diagnostic AD; the group *n* shown per panel are after independent Tukey 3×IQR outlier pruning (n = 961/141/86 for conductance and 961/133/81 for RCI). ADNI is an independent, 53-site cohort ^21, 22^; panels show the ComBat-harmonized full cohort (*n* = 368/209/32, i.e. the 368 of 372 CN with a computable global conductance, plus 209 MCI and 32 AD; see Methods) ^40, 41^. The CN-vs-AD conductance effect also survives, without harmonization, in the single largest homogeneous acquisition stratum (paired 2mm, *n* = 412; conductance defined for 245 of 247 CN). Replication AUCs for every contrast and stratum are tabulated in Supplementary Table S3. All four panels show the same monotone CN > MCI > AD gradient observed for RCI in Figure 2.

## Discussion

We developed two digital indices of periventricular white matter routing computed automatically from standard diffusion MRI: RCI, which scores how closely an individual’s routes match those used by a cognitively normal reference population, and a companion global conductance metric. We evaluated them across five stages of the AD trajectory and found that RCI was reduced in sessions acquired before disease conversion. RCI declined at every subsequent stage without reversal. Its degradation was anatomically structured rather than diffuse. A machine learning model built on these routing features, together with demographic, structural-MRI and cognitive covariates, identified individuals up to four years before their clinical conversion. Fluid biomarker assays, now in clinical use, indicate the presence of amyloid pathology. In contrast to blood-based biomarkers, changes in RCI may be reduced years before the onset of clinical symptoms and change earlier than atrophy measures.

RCI and global conductance were computed from the diffusion of water within tissue, which reports on the microstructure and orientation of white-matter pathways. Thus, they describe the structural configuration of those pathways rather than bulk fluid transport along them. Four independent lines of evidence argue that what they capture is nevertheless not simply generic neurodegeneration. First, degradation is anatomically concentrated: fronto-striatal routes degrade far more than the canonical hippocampal-fornix backbone (Figure 4), a pattern that would not be expected from diffuse atrophy alone. Fronto-striatal pathways connect the frontal lobe, which supports planning, personality, and executive control, to the basal ganglia, which govern action selection and habit; they subserve executive function, goal-directed behavior, reward processing, and motor control, and frontal-subcortical circuit disruption is an established substrate of the cognitive and behavioral symptoms seen in AD and related dementias ^16, 17^. Second, the signal converges with CSF amyloid-*β*_42_ in the expected direction (plasma pTau-217 only in a single-protocol stratum), while the continuous amyloid-PET and tau-PET associations were null. Third, allele dose at rs12454744 (located immediately upstream of AQP4 and within an astrocyte-specific super-enhancer identified in cell-type-resolved brain epigenomic maps ^15^, where AQP4 is the astrocytic water channel required for perivascular fluid transport ^18^) showed a direction-consistent, nominally significant dose-response relationship specifically with RCI (not conductance) in ADNI, while APOE4 copy number showed no dose-response with either metric; this metric-specific pattern is consistent with a genuine link to AQP4-mediated fluid transport rather than a generic genotype confound, though the AQP4 result did not survive multiple-comparison correction in either of the two strata tested and should be treated as hypothesis-generating rather than confirmatory. Fourth, a feature-ablation analysis showed that adding age, sex, and APOE4 to the structural-routing model did not improve discrimination (ROC-AUC 0.812 with versus 0.813 without), so their demographic information is already captured by the routing features. None of these observations is individually decisive, and none establishes that the signal is specific to fluid transport rather than to white-matter integrity more generally. Together they argue only that the measurement is not a restatement of atrophy, age, or genotype.

The inputs to both metrics, streamline counts, fractional anisotropy, and radial diffusivity, are themselves sensitive to axonal loss, demyelination, regional volume change, and partial-volume effects, all of which occur in AD, so some overlap with atrophy is expected. Several observations nonetheless suggest the routing signal is not simply a restatement of atrophy. Its degradation is anatomically selective, concentrating in fronto-striatal routes while the canonical hippocampal-fornix backbone, among the earliest and most severely atrophic territories in AD, is comparatively preserved, so the anatomy of greatest routing vulnerability does not track the anatomy of greatest atrophy. RCI is also the only marker examined whose five-group effect size increases rather than decreases after adjustment for age, sex, and APOE4 (*ε*^2^ from 0.166 to 0.193), whereas hippocampal volume (0.275 to 0.204) and the other markers decline. Together with the amyloid and AQP4 convergence noted above, this is consistent with a connectional signal at least partly distinct from generic atrophy, though the two cannot be fully separated with cross-sectional data.

Because the underlying metrics require only a standard clinical diffusion MRI sequence and are computed without manual input, this approach is, in principle, deployable on widely available clinical MRI scanners, although diffusion imaging is not yet part of standard dementia workup. It is not a replacement for amyloid PET or CSF testing, which establish molecular pathology directly; its likely role is complementary, as a lower-burden way to help prioritise which already-referred patients proceed to confirmatory molecular workup. Where a blood-based assay establishes that pathology is present, an imaging index of routing describes what has already changed in the white matter, and where. The weak correlation between RCI and plasma pTau-217 observed here suggests the two may carry additive information, which is directly testable in cohorts where both are available. Given the modest precision at fixed operating points noted above, the most defensible near-term role is risk stratification within an already-referred, higher-prevalence clinical population rather than unenriched population-wide screening, pending prospective validation.

The RCI and the global conductance estimator are, by construction, not AD-specific algorithms: both learn, from any labelled reference population, how a phenotype reshapes structural routing through the connectome, and then score an individual against that reference. The two differ in what they summarise, which may explain why they behave differently here: conductance is a single global scalar over the whole network, whereas RCI is referenced to a population and resolved at the level of individual paths. RCI declined monotonically; conductance did not. Whether the signal they capture is biologically specific to AD, as opposed to a more general marker of neurodegeneration, is not established by the present data; resolving this will require applying the same method across other conditions spanning the cognitive and disease spectrum. This has two practical consequences. First, because RCI collapses a high-dimensional routing pattern into a single, monotone, covariate-adjusted scalar, it is well suited as an engineered input feature or a pre-computed index for downstream machine-learning and disease-forecasting models, rather than only as a standalone test. Second, a continuous, imaging-only index that separates disease stages and anticipates conversion could be used for biomarker-based cohort enrichment in clinical trials, where selecting higher-risk or faster-progressing participants can materially reduce required sample size ^19^.

Because RCI is calibrated on labelled populations it is a supervised index, so a head-to-head with unsupervised scalar markers such as hippocampal volume or radial diffusivity is not strictly like-for-like. Two observations indicate that supervision is not what generates its performance. First, the concordance weights are estimated only from stable-CN and post-diagnostic sessions; the pre-clinical group that carries the headline result is never seen during fitting, so its discrimination is an out-of-sample property of the learned reference rather than a fit to the tested labels. Second, supervision with far more information does not do better: a gradient-boosted classifier trained on the full 2,306-feature connectome matrix reaches an out-of-fold AUC of 0.814 for pre-clinical conversion, no better than a classifier restricted to structural-routing features alone (0.813) or with age, sex, and APOE4 added (0.812) in a separate ablation run. It is therefore the routing structure the index summarises, not the act of supervision or the size of the feature space, that carries the signal. A fuller benchmark against other supervised connectome models, such as regularised edge classifiers, graph neural networks, or connectome-derived components, is a natural next step, but would test how best to read the routing signal rather than whether it exists.

Both discovery and replication cohorts are cross-sectional and retrospective; prospective, longitudinal designs with serial diffusion imaging are needed to establish predictive validity at the individual level. Negative (stable-CN) labels are only partially verified: 422 of the 991 stable-CN sessions (42.6%) had less than one year of subsequent follow-up, so a minority of apparent non-converters could convert beyond the observation window, which would inflate specificity. Under a stricter label that counts as negatives only stable-CN participants with at least four years of documented follow-up (plus converters whose conversion occurred more than four years after imaging), a model without cognitive covariates reaches an AUC of 0.780 [0.737, 0.823], versus 0.823 for the same model under the primary label; the signal is therefore attenuated but remains well above chance. ADNI’s PET, CSF and plasma sub-cohorts are small (*n* = 34–132) and were assayed a median of six to seven years from the diffusion acquisition, so those correlations should be read as cross-sectional structural associations rather than concurrent validation. Radial diffusivity and global conductance were numerically worse in pre-clinical AD than in either post-diagnostic group; we cannot at present distinguish whether this reflects selection into the post-diagnostic groups, differential survival, or differences in acquisition era. Finally, both cohorts under-represent non-white participants and were acquired in research rather than clinical settings, so generalisation to routine clinical populations remains to be demonstrated. We also cannot exclude that route degradation partly reflects vascular white-matter injury, although white-matter hyperintensity volume discriminated considerably less well than RCI (0.617 versus 0.817), arguing against a purely vascular explanation.

The decisive next steps are prospective, longitudinal cohorts with serial DTI to establish within-subject predictive validity rather than cross-sectional separation, and pairing RCI with plasma biomarkers such as pTau-217, with which it correlated only partially here, to test whether the two carry additive information; and application of the same construction across other conditions, to determine whether the signal is specific to AD or reflects white-matter vulnerability more generally. Because these metrics are computed from a standard diffusion sequence acquired alongside structural imaging in most protocols, and because that data is at present largely unused in the workup of dementia, the imaging required to pursue these questions has in many cases already been obtained.

## Methods

### Participants

OASIS-3 is a longitudinal, single-site cohort of cognitively normal and AD participants aged 42–95 years ^20^. After quality control, 1,706 MRI sessions from 1,027 unique participants were retained. Of these, 1,656 were assignable to one of five mutually exclusive trajectory groups based on longitudinal clinical follow-up: stable CN (*n* = 991), pre-clinical MCI (*n* = 230), pre-clinical AD (*n* = 205), post-diagnostic MCI (*n* = 144), and post-diagnostic AD (*n* = 86); the remaining 50 sessions had an indeterminate trajectory (insufficient follow-up to classify) and were excluded from group-wise analyses. Trajectory groups were defined from longitudinal Clinical Dementia Rating (CDR) scores: conversion to MCI was taken as the first visit reaching CDR ≥ 0.5 and conversion to dementia as CDR ≥ 1.0. Pre-clinical MCI and pre-clinical AD sessions were scored at CDR = 0 (or ≤ 0.5 for pre-AD) at the imaging visit but converted at a later visit; post-diagnostic sessions were imaged after a documented conversion; stable CN sessions had CDR = 0 with no recorded subsequent conversion. ADNI is a longitudinal, 53-site, multi-protocol cohort ^21, 22^. The primary ADNI replication analyses use a ComBat-harmonized cohort of *n* = 609 sessions (368 CN, 209 MCI, 32 AD): every session with a usable diagnostic label and a computable harmonized global-conductance/RCI value. This is the sample plotted in Figure 7 and quantified in Supplementary Table S3, and the *n* we report throughout. (It derives from 686 imaged ADNI sessions, of which 372 CN, 209 MCI, and 32 AD carried a diagnostic label; four CN lacked a computable global conductance and are excluded, giving the 368 CN above.) A single-protocol (paired 2mm, *n* = 412: 247 CN, 144 MCI, 21 AD; conductance computable for 245 of the 247 CN) stratum is reported alongside as a scanner-confound check. Full imaging inclusion/exclusion criteria and Institutional Review Board approvals are described in the original cohort publications; the trajectory-group definitions above are specific to this study.

### Imaging acquisition and connectome construction

Diffusion-weighted images were processed to obtain voxel-wise fractional anisotropy, radial and axial diffusivity, and the primary diffusion eigenvector from the diffusion tensor ^23, 24^, then parcellated into 80 regions using a combined Desikan–Killiany–Tourville/FreeSurfer atlas ^25, 26, 27^. Whole-brain deterministic tractography ^28^ produced an 80×80 structural connectivity matrix (streamline count, mean FA, mean RD per region pair) per session. Four periventricular regions (bilateral thalamus, bilateral caudate) were designated source nodes (candidate sites of perivascular CSF entry) and paired with each of the remaining 79 regions, yielding 316 source→target routes per session.

OASIS-3 diffusion data are heterogeneous across sessions, comprising single-shell DTI, multi-shell, and diffusion-spectrum-imaging (DSI) acquisition schemes accrued over the cohort’s collection period. All sessions were reconstructed with a single generalized *q*-sampling imaging pipeline (DSI-Studio ^29^) on the identical 80-region atlas, so every downstream routing metric is computed uniformly regardless of acquisition, and residual acquisition differences are further mitigated by within-cohort normative *z*-scoring. We confirm in Results that the biomarker’s discrimination is stable within each acquisition scheme.

### Structural routing metrics

Full mathematical derivation and formal justification of both metrics, including a worked proof of the effective-conductance circuit interpretation, are provided in the Supplementary Methods. Briefly: RCI applies Yen’s *K*-shortest-paths algorithm (*K* = 50) ^30^, built on Dijkstra’s algorithm ^31^, to enumerate the most structurally favorable routes between each source and target, then scores a session’s observed path usage against log-likelihood-ratio weights estimated from a held-out, leakage-controlled training-fold CN/disease split. The score is positive when routing resembles the CN reference population and negative when it resembles the disease reference population, modulated by each path’s structural quality. Critically, the pre-clinical (pre-MCI and pre-AD) sessions are excluded from weight estimation and enter only as held-out test cases, so the pre-clinical discrimination reported below is not circular. Global conductance converts the 80 × 80 connectivity matrix into a weighted graph, 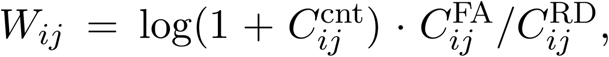 computes the graph Laplacian **L** = **D** − **W**, and obtains the Kirchhoff effective conductance between any source-target pair from the Moore–Penrose pseudoinverse **L**^+^ ^32, 33^, following established graph-theoretic treatments of brain connectivity ^34^. The reported scalar biomarker, which we call global conductance, is the mean of this per-pair effective conductance over all 316 routes. For comparison with the established glymphatic-MRI proxy, a conventional DTI-ALPS index ^11^ was also computed as a local measure of perivascular diffusivity adjacent to the lateral ventricle.

Algorithms 1 and 2 summarize both procedures at implementation level; the corresponding closed-form expressions, the Naive Bayes derivation of Algorithm 1, and the Kirchhoff circuit proof of Algorithm 2 are given in the Supplementary Methods.

#### Algorithm 1 RCI: population calibration and per-session scoring

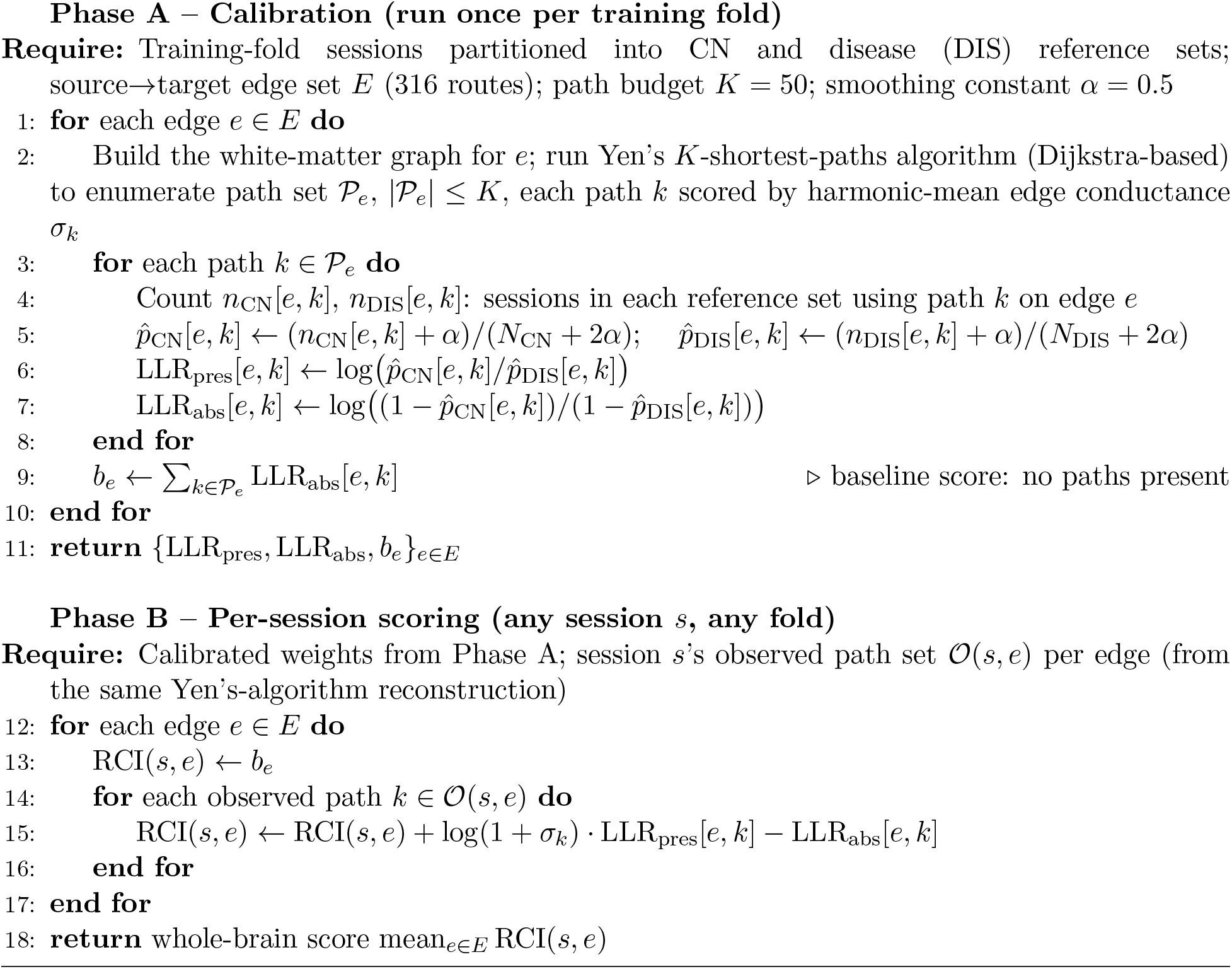

#### Algorithm 2 Effective conductance via graph-Laplacian pseudoinverse

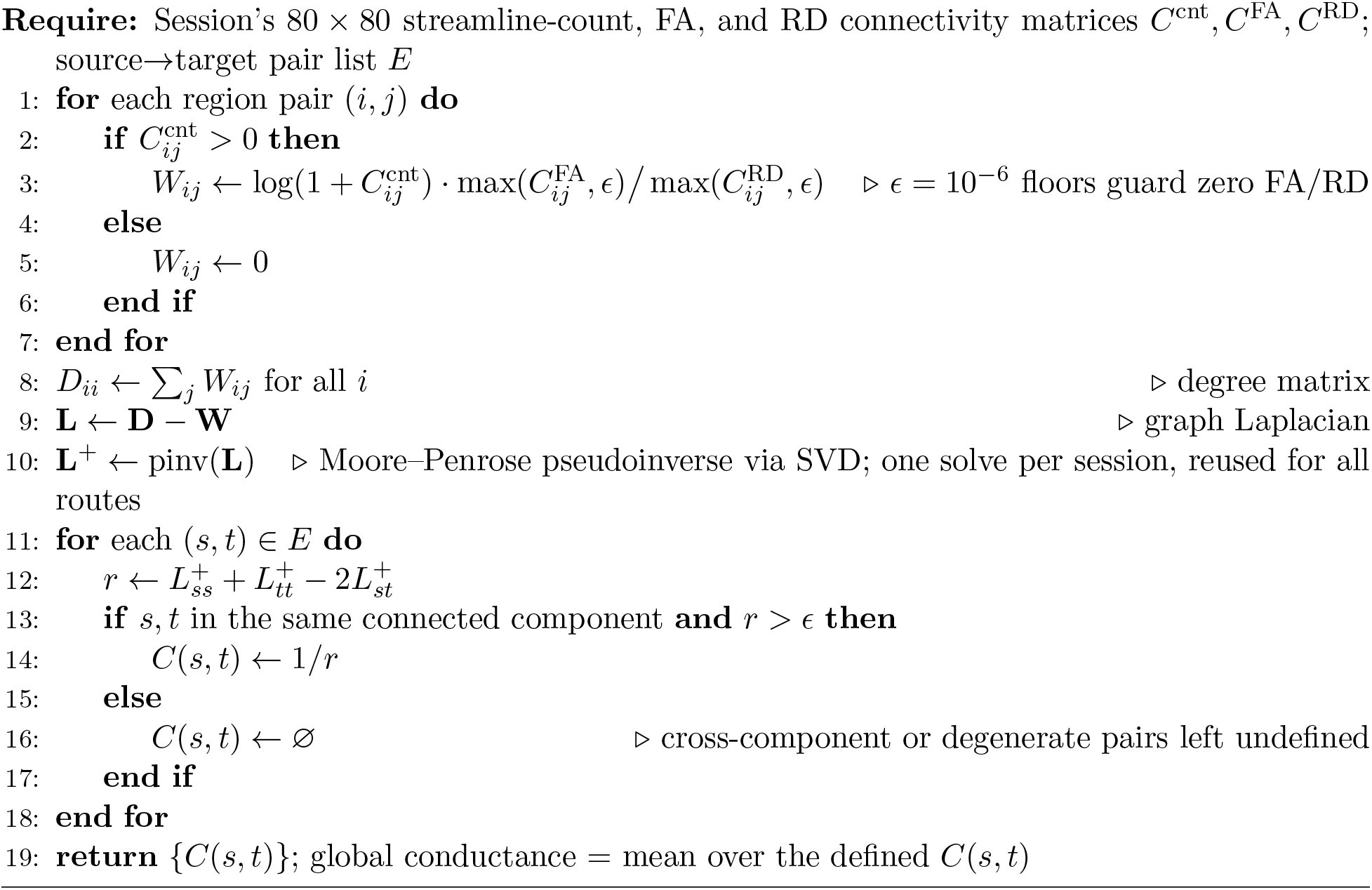

### A network-level pipeline for inferring white-matter routing integrity from clinical DTI

Figure 1 and Supplementary Fig. S1 summarize the two metrics used throughout this paper. Effective conductance (Figure 1) treats the structural connectome as a resistor network and computes, via the Kirchhoff circuit laws encoded in the graph Laplacian’s pseudoinverse, the total current that would flow between a source and target region under a unit voltage, a quantity that increases with every redundant parallel pathway and decreases when multiple parallel routes are simultaneously degraded. RCI (Figure S1) instead asks how closely a participant’s actual, observed white-matter routes match the routes preferentially used by a cognitively normal reference population, weighted by each route’s structural quality. Both metrics are computed on the same 316 periventricular-source → target routes, are derived independently (one is a global network solve, the other a population log-likelihood score over enumerated paths), and, as shown below, yield convergent disease gradients despite being algorithmically unrelated, which constitutes a form of mutual validation.

### Machine learning and statistics

Our contribution is the structural-routing feature set itself (RCI, global conductance, and their constituent edge-level features, Algorithms 1–2); to evaluate its predictive utility as a biomarker we passed it into a standard, off-the-shelf classifier rather than a bespoke one. Preclinical detection used a histogram-based gradient-boosted classifier (implemented via the scikit-learn library ^35^) under 25-fold repeated, subject-grouped, stratified cross-validation, with within-training-fold SMOTE class rebalancing ^36^ and fold-specific re-estimation of RCI log-likelihood weights to prevent leakage. An ordinal extension used a Frank–Hall threshold decomposition ^13^. Feature importance was assessed with SHAP ^37^. Group comparisons used Kruskal–Wallis and Mann–Whitney *U* tests with Bonferroni correction; classifier discrimination used DeLong’s method for AUC confidence intervals ^38^. Because competing biomarkers are measured on the same sessions, head-to-head AUC differences were tested with DeLong’s paired test for two correlated ROC curves ^38^, restricted to sessions with both measures present; we report ΔAUC with a 95% CI and a two-sided *p* (Supplementary Table S2). Age/sex propensity-score matching followed standard caliper-width guidance ^39^. ADNI multi-protocol harmonization used ComBat ^40^, following established application to diffusion MRI ^41^. Full statistical protocols and the complete propensity-matched comparison table, the full ADNI biological-validation tables, and the cross-validation and hyperparameter details are provided in the Supplementary Methods.

## Data Availability

All data produced are available online at

https://www.oasis-brains.org

https://adni.loni.usc.edu

## Data availability

OASIS-3 and ADNI imaging and clinical data are available to qualified investigators from their respective data-sharing platforms (OASIS-3: https://www.oasis-brains.org; ADNI: https://adni.loni.usc.edu) subject to each study’s data-use agreement. No participant-level data are redistributed by this work. The group-level aggregate result tables underlying the main figures are provided in the analysis repository (see Code availability).

## Code availability

All analysis code and the group-level aggregate results needed to regenerate the main figures are openly available under the MIT license at https://github.com/mergho28/rci-alzheimers-analysis. A standalone, pip-installable implementation of the global-conductance and Route Concordance Index metrics (connectome-rci) is available at https://github.com/mergho28/connectome-rci.

## Acknowledgements

Data used in preparation of this article were obtained in part from the Alzheimer’s Disease Neuroimaging Initiative (ADNI) database (adni.loni.usc.edu). As such, the investigators within the ADNI contributed to the design and implementation of ADNI and/or provided data but did not participate in analysis or writing of this report. A complete listing of ADNI investigators can be found at http://adni.loni.usc.edu/wp-content/uploads/how_to_apply/ADNI_Acknowledgement_List.pdf.

ADNI is funded by the National Institute on Aging (National Institutes of Health Grant U01 AG024904) and the Department of Defense (award W81XWH-12-2-0012), with contributions from the private-sector partners listed in the ADNI Data Use Agreement.

Data were provided in part by OASIS-3 (Longitudinal Multimodal Neuroimaging; Principal Investigators T. Benzinger, D. Marcus, J. Morris), supported by National Institutes of Health grants P30 AG066444, P50 AG00561, P30 NS09857781, P01 AG026276, P01 AG003991, R01 AG043434, UL1 TR000448, and R01 EB009352.

The work is supported by UCSD Shiley-Marcos Alzheimer’s Disease Research Center (P30 AG062429).

## Author contributions

M.G. conceived the study, developed the methods, performed the analyses and wrote the manuscript; J.C.M.S. supervised the work and revised the manuscript. All authors approved the final version.

## Supplementary Methods

**Figure S1:**
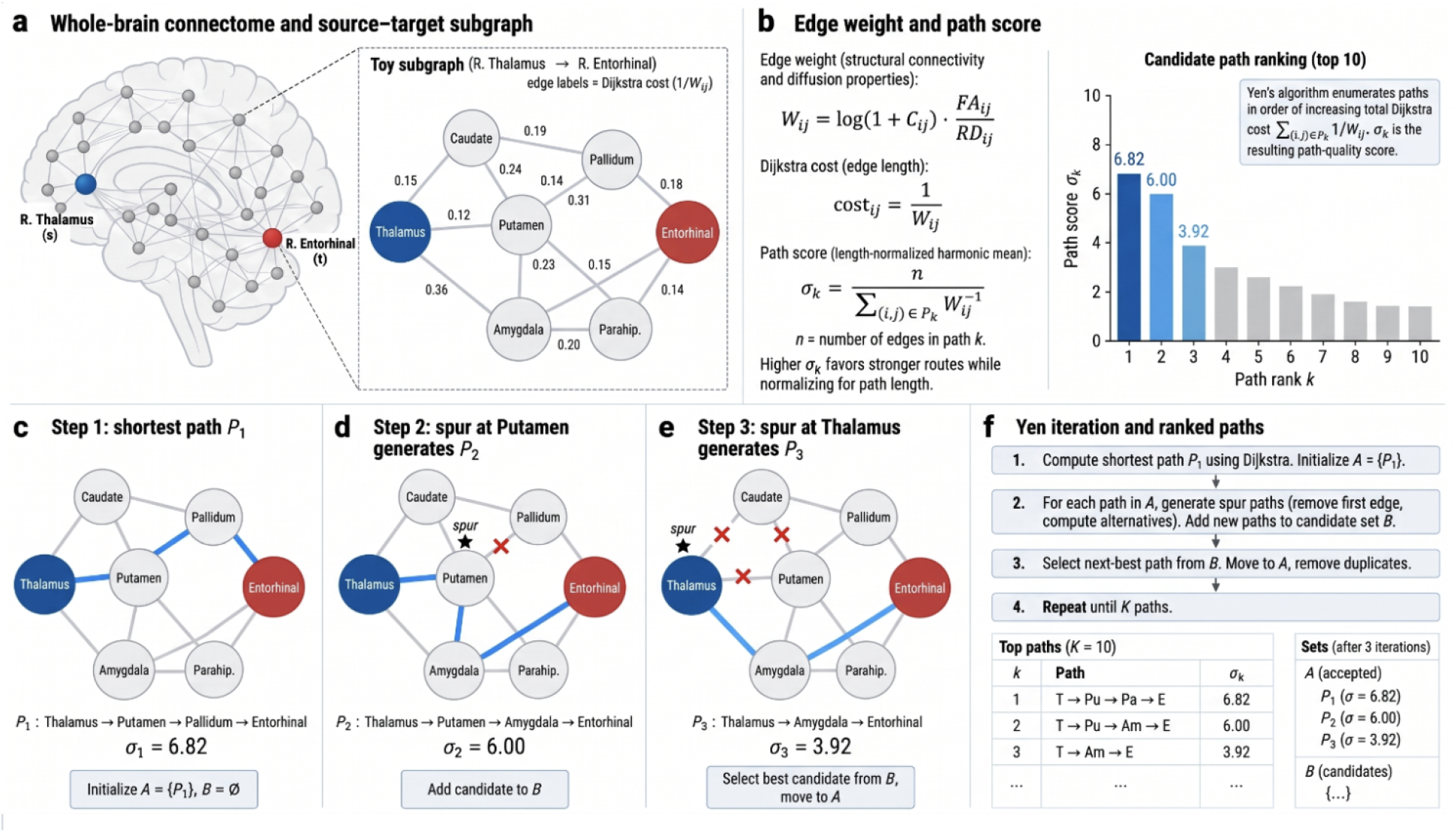
Route enumeration underlying the Route Concordance Index (RCI). Yen’s *K*-shortest-paths algorithm ^30^ (built on Dijkstra’s shortest-path algorithm ^31^) enumerates the *K* = 50 structurally best routes between each periventricular source and cortical/subcortical target, ranked by harmonic-mean edge conductance. These ranked path inventories are scored against log-likelihood-ratio weights learned from a cognitively normal reference population to yield the path-concordance (RCI) score, fully derived in the Supplementary Methods. The routes are the same 316 periventricular source→target pairs defined in Figure 1.

This appendix provides the complete mathematical derivation, formal justification, and extended statistical tables supporting the main text above. It motivates the RCI path-scoring algorithm (Algorithm 1) from a Naive Bayes log-odds framework, proves the baseline-plus-evidence decomposition for the binary case, and states explicitly where the quality-weighted score departs from it; proves the Kirchhoff circuit interpretation of the graph-Laplacian effective-conductance estimator (Algorithm 2) and establishes its key monotonicity property; details the normative z-scoring and residualization protocol; gives the full propensity-score-matching protocol and results table; reports the complete ADNI external-validation statistical tables (Tier A replication AUCs and Tier B biological-specificity correlations) summarized in the main text; and details cross-validation, model hyperparameters, and leakage-control procedures.

**Figure S2:**
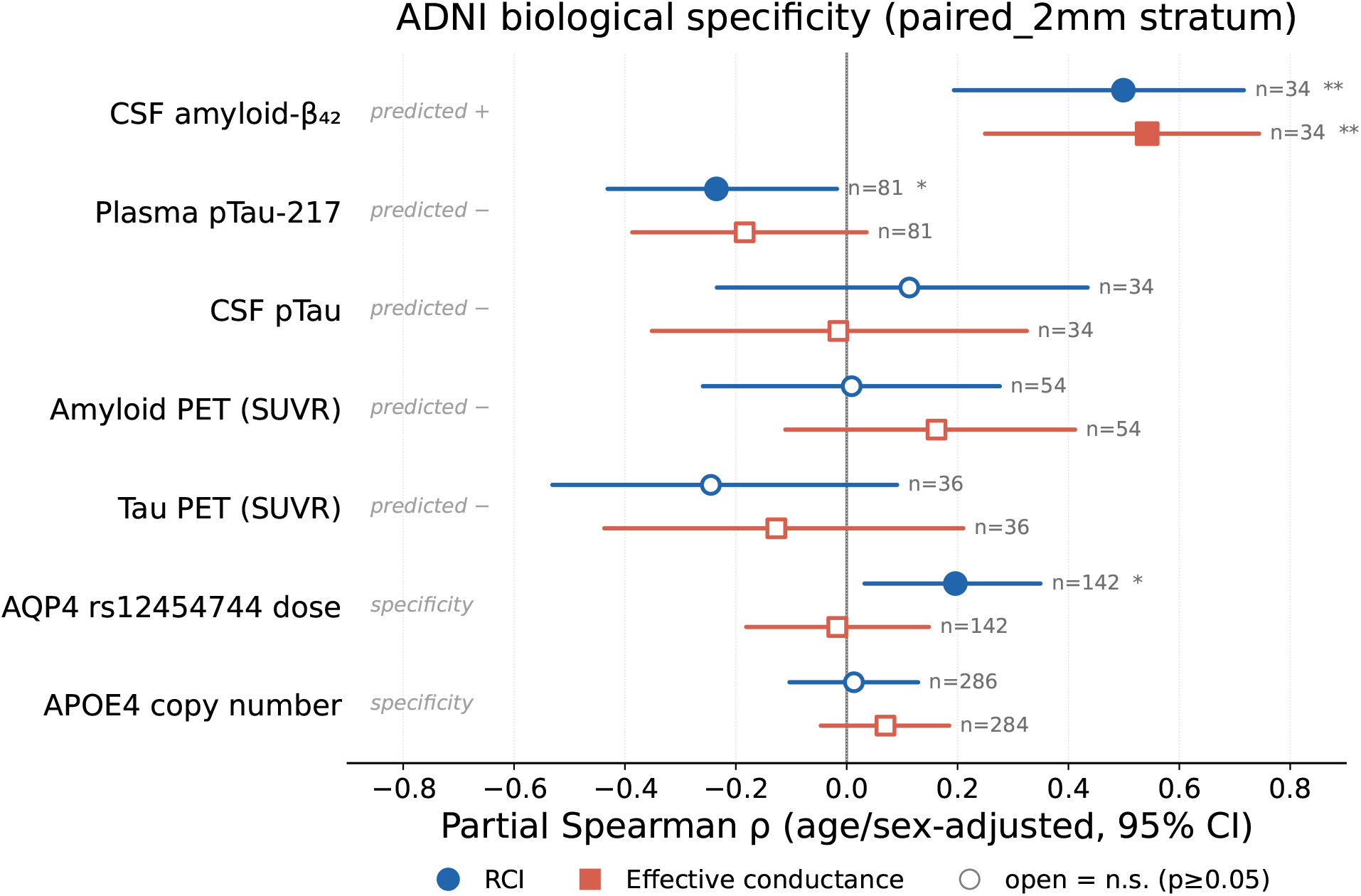
Biological specificity of RCI and global conductance in ADNI. Age/sex-adjusted partial Spearman correlations (95% CI, Fisher-*z*) of each metric with molecular AD markers and genotype dose, all in the single-protocol paired 2mm stratum (no harmonization). Each row uses the subset of paired 2mm participants who had both a usable DTI session and that assay, so *n* varies by row and is shown per marker: CSF markers require a lumbar puncture (*n* = 34) and PET a dedicated scan (*n* = 36–54), so these sub-samples are small, whereas plasma (*n* = 81) and genotype (*n* = 142–286) are available for more participants; the wide confidence intervals on the small-*n* rows reflect this limited power. Filled markers are significant (*p* < 0.05), open markers not; italic tags give the direction predicted under the fluid-clearance hypothesis. CSF amyloid-*β*_42_ is the one marker associated with both metrics in the predicted direction (RCI *ρ* = 0.50, conductance *ρ* = 0.54; both *p* < 0.01); plasma pTau-217 correlated with RCI in this single-protocol stratum (*ρ* = −0.24, *p* = 0.035), an association that did not persist in the ComBat-harmonized full cohort (Supplementary Table S4) and is therefore not treated as RCI-specific. Amyloid PET, tau PET, CSF pTau, and APOE4 dose are null for both metrics, and AQP4 rs12454744 dose shows the RCI-specific (*ρ* = 0.20, *p* = 0.020), conductance-null pattern discussed in the text. All correlations are uncorrected for the 14 comparisons in this panel and are hypothesis-generating (full values, and the ComBat-harmonized full-cohort estimates, in Supplementary Table S4).

## RCI: A Naive-Bayes-Motivated Path-Concordance Score

### Biological motivation and path inventory

The glymphatic system preferentially routes cerebrospinal fluid (CSF) along white-matter perivascular spaces. We hypothesize that cognitively normal (CN) brains traverse reproducible, stereotyped white-matter routes from periventricular CSF sources to cortical targets, and that these preferred routes are progressively disrupted across the Alzheimer’s disease (AD) trajectory. RCI quantifies how closely a participant’s observed streamline routing resembles that of a healthy reference population.

For each session *s* and each of the 316 source→target pairs *e*, the *K* = 50 shortest paths through the weighted white-matter graph are enumerated using Yen’s *K*-shortest-paths algorithm ^30^, which iteratively applies Dijkstra’s shortest-path algorithm ^31^ with edge removal to generate successive loopless alternatives (Algorithm 1, Phase A). Each path *k* is a sequence of atlas-region indices from source to target, scored by its harmonic-mean edge conductance *σ_k_* > 0 (higher = structurally stronger).

### Prevalence estimation with Jeffreys-prior smoothing

Let *P* denote the set of all unique paths observed on edge *e* across the training cohort. For path *k* ∈ *P*, prevalence in the CN and disease (DIS) reference populations is estimated with Jeffreys-prior Laplace smoothing (*α* = 0.5):

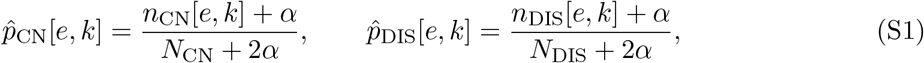

where *n*_CN_[*e*, *k*] is the number of CN sessions using path *k* on edge *e*, and *N*_CN_ the total number of CN sessions (similarly for DIS). The CN reference comprises all stable-normal training-fold sessions; the disease reference comprises all post-MCI and post-AD training-fold sessions. Pre-MCI and pre-AD sessions are excluded from this fitting step to prevent label leakage into the pre-clinical prediction target evaluated in the main-text machine-learning analysis (Figure 6).

#### Jeffreys prior justification

For a binomial observation with *n* successes out of *N* trials, the Jeffreys prior ^42^ is Beta(1/2, 1/2), yielding posterior-mean estimate *p̂* = (*n* + 1/2)/(*N* + 1). We use the equivalent Laplace-smoothed form (*n* + *α*)/(*N* + 2*α*) with *α* = 0.5, which prevents log(0) singularities when a path is unobserved in one cohort while retaining asymptotic consistency as *N* → *∞*.

#### Log-likelihood-ratio weights

For each path k on edge e, two log-likelihood-ratio (LLR) weights are computed:

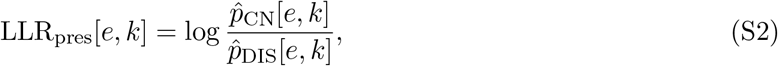

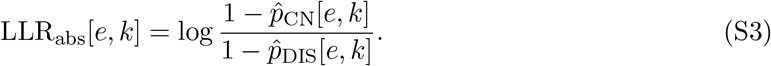

A positive LLR_pres_ indicates a path more prevalent in CN than disease (the subject is rewarded for using it); a negative LLR_abs_ indicates that not using a CN-typical path is itself evidence of disease (the subject is penalized for its absence).

### Formal derivation: RCI as a Naive Bayes log-posterior-odds score

#### Proposition 1 (Naive Bayes path-evidence decomposition)

*Consider binary classification (CN vs. disease) where the observation for edge e is the multiset of paths O*(*s*, *e*) = {*k*_1_, *k*_2_, …} *used by session s. Assume conditional independence of path usage across k* ∈ *P given class membership (the Naive Bayes assumption). Then the log-posterior odds of CN over disease decompose as a sum of per-path evidence terms*.

*Proof.* Let *x_k_* ∈ {0, 1} indicate whether session *s* used path *k* on edge *e*. Under conditional independence,

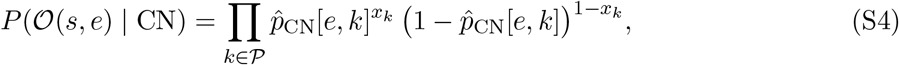

and likewise for disease with *p̂*_DIS_. By Bayes’ rule,

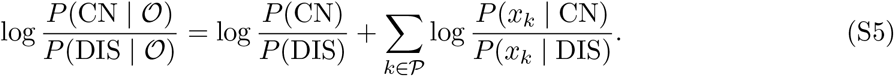

Each summand expands as

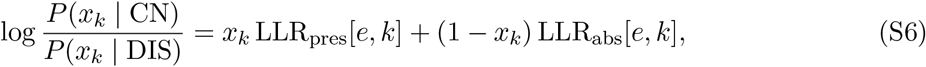

using definitions (S2)–(S3). Dropping the class-prior term (equal priors assumed) and separating the sum over observed (*x_k_* = 1) versus unobserved (*x_k_* = 0) paths,

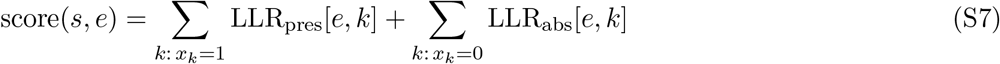

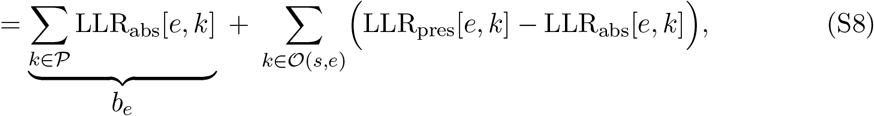

where the second line follows by adding and subtracting Σ*_k_*_∈*O*(*s,e*)_ LLR_abs_[*e, k*] and noting that the sum over *k* ∈/ *O*(*s, e*) of LLR_abs_ combines with the sum over *k* ∈ *O*(*s, e*) of LLR_abs_ to recover the full-path-set sum *b_e_*. This establishes the baseline-plus-evidence decomposition used throughout.

### Quality-weighted extension and the closed-form RCI score

Equation (S8) treats path usage as a binary indicator. RCI extends this by replacing the indicator *x_k_* = 1 with a continuous, quality-weighted contribution log(1 + *σ_k_*), so that structurally strong paths (high harmonic-mean conductance *σ_k_*) contribute more evidence than structurally weak paths at the same anatomical route (Algorithm 1, Phase B). The resulting score for session *s* on edge *e* is

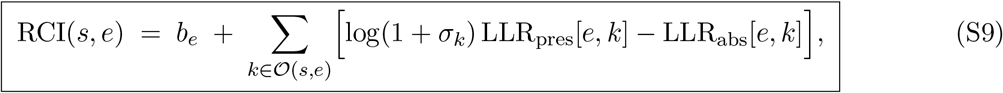

with *b_e_* = Σ*_k_*_∈*P*_ LLR_abs_[*e*, *k*] as in Proposition 1. The per-session, whole-brain RCI biomarker score reported in the main text is the mean of RCI(*s, e*) over all 316 edges *e*. Higher values indicate a CN-like routing signature; negative values indicate a disease-like signature.

We state plainly that this quality weighting is a heuristic extension, not a consequence of Proposition 1. The Proposition establishes the baseline-plus-evidence decomposition for the binary indicator; multiplying LLR_pres_ by log(1 + *σ_k_*) departs from that derivation, so Equation (S9) is

Naive-Bayes-motivated rather than a calibrated log-posterior-odds, and RCI should not be read as a posterior probability. The weighting is applied to the presence term only, leaving the absence penalty LLR_abs_ unweighted; Equation (S9) reduces exactly to the Naive Bayes form of Equation (S8) when log(1 + *σ_k_*) = 1. We adopt it because path quality is informative beyond mere presence, and we validate the resulting score empirically rather than by appeal to the Naive Bayes derivation.

### Leakage control

In cross-validation, the LLR weights in Equations (S2)–(S3) are re-estimated from scratch using only training-fold CN and post-diagnostic sessions in every fold, so that no test-session path pattern influences the weights used to score that session. Without this correction, cross-validated AUC on the binary pre-clinical task was inflated by approximately 0.03 (Pearson *r* = 0.976 between leakage-corrected and uncorrected scores), confirming both that the correction matters and that the corrected and uncorrected scores are highly concordant in rank. All AUC/PR-AUC values reported in the main text (including Table 1 and Figure 6) use leakage-corrected out-of-fold predictions.

## Effective Conductance: Kirchhoff Circuit Interpretation and Proof

### Edge weights

The 80 × 80 structural connectivity matrix is converted to an edge-weight matrix **W** combining streamline count, fractional anisotropy (FA), and radial diffusivity (RD):

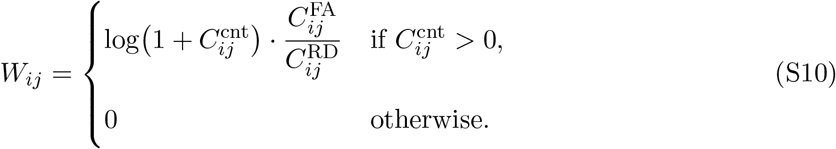

This rewards connections supported by many streamlines, with high FA (intact myelin) and low RD (intact axonal membranes) ^43^.

### Graph Laplacian and its pseudoinverse

Treating **W** as a resistor network in which each edge is a conductor of weight W*_ij_*, the graph Laplacian is

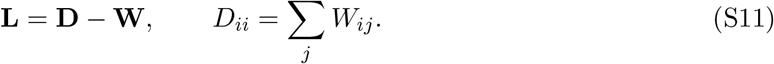

**L** is symmetric, positive semi-definite, and singular (every row sums to zero, so **1** is in its null space), hence not directly invertible. Its Moore–Penrose pseudoinverse **L**^+^ is obtained via the singular value decomposition **L** = **UΣU***^⊤^* as **L**^+^ = **UΣ**^+^**U***^⊤^*, inverting each non-zero eigenvalue (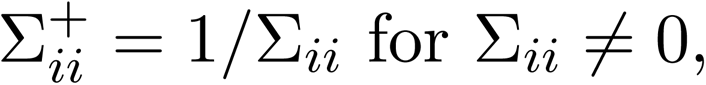 skipping the zero eigenvalue associated with **1**); see Algorithm 2.

### Theorem: effective resistance from the Laplacian pseudoinverse

#### Theorem 1 (Effective resistance, after Klein & Randić ^32, 33^).

Let **L**^+^ be the Moore–Penrose pseudoinverse of the weighted graph Laplacian **L** for a connected resistor network with conductances *W_ij_. The effective resistance between nodes s and t is*

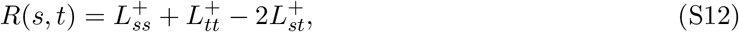

*and the effective conductance is* C(s, t) = 1/R(s, t).

*Proof.* Inject a unit current at node *s* and extract it at node *t*; let **b** = **e***_s_* − **e***_t_* be the corresponding current-injection vector, where **e***_i_* is the i-th standard basis vector. Kirchhoff’s current law states that the nodal voltage vector **v** satisfies **Lv** = **b**. Because **b** ⊥ **1** (it sums to zero) and **1** spans the null space of **L**, this system has the minimum-norm solution

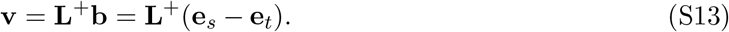

By Ohm’s law, the effective resistance is the voltage drop required to drive a unit current from *s* to *t*:

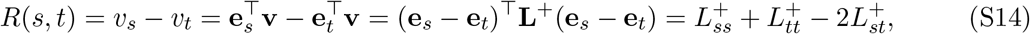

using the symmetry of **L**^+^. Since **L** is positive semi-definite with null space spanned only by **1** on a connected graph, this quadratic form is strictly positive for *s* ≠ *t*, so *R*(*s, t*) > 0 and *C*(*s, t*) = 1/*R*(*s, t*) is well defined. ▪

### Lemma: monotonicity under parallel-pathway addition

#### Lemma 1 (Conductance increases with parallel pathways).

Let **W** and **W**^′^ = **W** + Δ *be two weighted adjacency matrices on the same node set, where* Δ ⪰ 0 *entrywise represents one or more additional or strengthened edges (additional parallel white-matter pathways). Then for any s* ≠ *t, C^′^*(*s, t*) ≥ *C*(*s, t*)*, i.e. effective conductance is monotonically non-decreasing in edge weight*.

*Proof.* This is a standard monotonicity property of effective resistance in resistor networks (Rayleigh’s monotonicity law): adding a parallel conductor between any pair of nodes, or increasing any existing conductance, cannot increase the effective resistance between any two terminals, because the additional conductance provides an additional current path that can only divert current away from existing paths, weakly reducing total dissipation for a fixed terminal voltage. Formally, 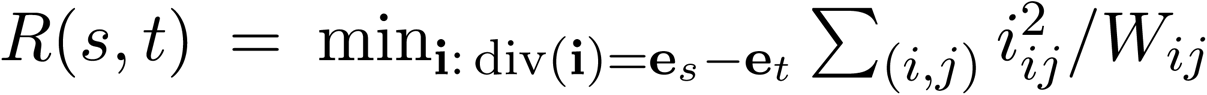 (Thomson’s principle: the physical current distribution minimizes power dissipation among all valid flows). Since 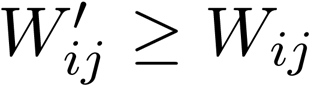 for all edges, the dissipation of any fixed admissible flow under **W***^′^* is no greater than under **W**, so the minimum over admissible flows cannot increase: *R^′^*(*s, t*) *≤ R*(*s, t*), hence *C^′^*(*s, t*) = 1/*R^′^*(*s, t*) ≥ 1/*R*(*s, t*) = *C*(*s, t*). ▪ □

#### Corollary 1.

Conversely, if k *of* n *parallel white-matter bundles connecting* s *and* t *are degraded (their weights reduced toward zero), the effective conductance* C(s, t) *is monotonically non-increasing in the number and severity of degraded bundles, and reflects the combined structural integrity of all* n *pathways rather than only the single strongest remaining pathway*.

This property is the central motivation for using effective conductance, rather than a single-shortest-path or single-ROI diffusivity metric, as a structural-routing biomarker: it is sensitive to distributed, partial degradation across a redundant network, which a shortest-path or single-region statistic is not.

### Conductance to AD-target regions

Connectivity to the eight Braak-stage I/III memory regions (bilateral entorhinal, parahippocampal, hippocampal, and amygdaloid cortices) is summarized as the mean conductance across the 32 source→AD-target pairs, 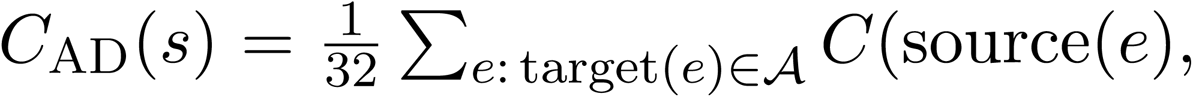 target(e)), where A is the set of eight AD-target region indices.

## Normative Z-Scoring Protocol

All edge-level features are normalized to age-, sex-, APOE4-, and FA-adjusted reference values derived from the CN population within each cross-validation training fold. For conductance, streamline count, RD, and mean traversal time, a log-transform is applied first, *x̃*(*s, e*) = log x(*s, e*) for x(*s, e*) > 0 (zero-valued edges are assigned the low-diffusivity sentinel described below rather than log-transformed), and an OLS model is fitted on training-fold CN sessions:

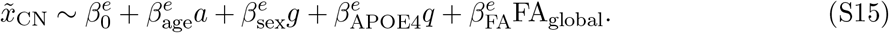

The normative z-score for session s is

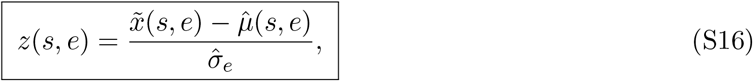

where *µ̂*(*s*, *e*) is the model-predicted value for session s’s demographics and *σ̂_e_* the residual SD from training-fold CN sessions on edge e. For FA and RCI, the same procedure omits the FA covariate. Sessions with no streamlines on edge e are assigned the sentinel value *z* = −3.0, except RCI, which incorporates an absence penalty analytically via Equation (S9). The normative model is always fitted using only training-fold CN sessions; test sessions are scored using training-fold parameters, preventing leakage.

## Propensity-Score Matching: Protocol and Full Results

To verify that group differences in structural-routing features are not driven by demographic confounds, we performed 1:1 nearest-neighbor propensity-score matching (PSM) on age and sex for each pairwise comparison (CN vs. pre-MCI, CN vs. pre-AD, CN vs. post-MCI, CN vs. post-AD). A logistic regression model estimated P(disease | age, sex); matched pairs were formed within a caliper of 0.2 × SD(*p̂*) using nearest-neighbor matching without replacement, following standard caliper-width guidance ^39^. Matched-pair counts: CN vs. pre-MCI (n = 230), CN vs. pre-AD (n = 196), CN vs. post-MCI (n = 142), CN vs. post-AD (n = 86). Because age and sex were combined into a single propensity score rather than exact-matched, this procedure balances the joint score but does not guarantee exact sex balance within matched pairs; residual sex imbalance is therefore a limitation of this analysis. Between-group differences in matched pairs were assessed by the Mann–Whitney U test; rank-biserial correlation r = 1 − 2U/(n_1_n_2_) is reported. Significance should be judged against a Bonferroni-corrected threshold for the full family of 28 tests (7 features × 4 contrasts), α = 0.05/28 ≈ 0.0018; the asterisks in Table S1 denote conventional uncorrected thresholds.

**Table S1:** Age/sex propensity-matched group comparisons (1:1 PSM, Mann–Whitney *U*). *r* = rank-biserial correlation; ns = not significant. Asterisks denote conventional uncorrected thresholds (∗ ∗ ∗∗ *p* < 0.0001, ∗∗ *p* < 0.01, ∗ *p* < 0.05); significance should be judged against the Bonferroni-corrected *α* = 0.05/28 ≈ 0.0018 for the 28-test family (see text).

| Feature | CN vs. pre-MCI | CN vs. pre-AD | CN vs. post-MCI | CN vs. post-AD |
| --- | --- | --- | --- | --- |
| RCI (LLR) | ns $r = -0.07$ | * ** * $r = -0.37$ | * ** * $r = -0.61$ | * ** * $r = -0.82$ |
| Conductance (AD targets) | ns $r = -0.06$ | * ** * $r = -0.28$ | ** $r = -0.18$ | ** $r = -0.29$ |
| Conductance (global) | ns $r = -0.05$ | * ** * $r = -0.30$ | ** $r = -0.19$ | ** $r = -0.28$ |
| Structural routes (count) | ns $r = +0.01$ | * ** * $r = -0.24$ | ns $r = -0.13$ | ** $r = -0.28$ |
| Radial diffusivity (316-route mean, unadjusted) | * $r = +0.12$ | ns $r = +0.04$ | ns $r = +0.05$ | ns $r = +0.02$ |
| Route entropy | ns | ns | ns | ns |
| Mean traversal time | ns | ns | ns | ns |

RCI is the strongest surviving biomarker after matching (r = −0.37 for CN vs. pre-AD, r = −0.82 for CN vs. post-AD; both p < 0.0001). Both global and AD-target-specific conductance remain significant for CN vs. pre-AD and CN vs. post-AD (all p < 0.05, uncorrected), as does the number of structural routes for CN vs. pre-AD (r = −0.24, p < 0.001). Route entropy and mean traversal time are not significant in any matched comparison, indicating these measures primarily reflect normal ageing rather than disease-specific routing disruption. No feature survives matching for the CN vs. pre-MCI comparison, consistent with the biological ambiguity of this transitional group.

## Paired DeLong Tests: Head-to-Head AUC Differences

Competing biomarkers are measured on the same sessions, so their ROC curves are correlated and the difference between two AUCs cannot be judged from overlapping marginal confidence intervals. We therefore tested each head-to-head difference with DeLong’s paired test for two correlated ROC curves ^38^, using the same age/sex/APOE4-residualized values and the same AUC orientation (P(CN-like group > disease-like group)) as Figure 3. Each test is restricted to the sessions with both measures present, which is why n differs slightly between rows (hippocampal volume and entorhinal thickness have some missing FreeSurfer values).

**Table S2:** Paired DeLong tests of ΔAUC (RCI minus comparator) on age/sex/APOE4-residualized biomarkers. Positive Δ favours RCI. *p* values are two-sided and uncorrected.

| Task | Comparator | $n$ (CN-like + disease-like) | $\text{AUC}_{\text{RCI}}$ | $\text{AUC}_{\text{comp}}$ | $\Delta\text{AUC}$ [95% CI] | $p$ |
| --- | --- | --- | --- | --- | --- | --- |
| CN vs. pre-AD | Hippocampal volume | 975+194 | 0.817 | 0.804 | +0.014 [−0.025, +0.053] | 0.49 |
| CN vs. pre-AD | Entorhinal thickness | 975+194 | 0.817 | 0.687 | +0.131 [+0.081, +0.180] | $2.5 \times 10^{-7}$ |
| CN vs. pre-AD | Global conductance | 991+205 | 0.817 | 0.713 | +0.104 [+0.059, +0.150] | $7.5 \times 10^{-6}$ |
| CN vs. pre-AD | Radial diffusivity | 991+205 | 0.817 | 0.822 | −0.005 [−0.045, +0.035] | 0.81 |
| CN vs. pre-AD | DTI-ALPS | 938+185 | 0.819 | 0.518 | +0.301 [+0.246, +0.356] | $< 10^{-26}$ |
| pre-MCI vs. post-MCI | Hippocampal volume | 222+138 | 0.634 | 0.667 | −0.033 [−0.095, +0.028] | 0.29 |
| pre-MCI vs. post-MCI | Entorhinal thickness | 222+138 | 0.634 | 0.606 | +0.028 [−0.051, +0.106] | 0.49 |
| pre-MCI vs. post-MCI | Global conductance | 230+144 | 0.638 | 0.544 | +0.094 [+0.012, +0.177] | 0.025 |
| pre-AD vs. post-AD | Hippocampal volume | 194+73 | 0.516 | 0.709 | −0.193 [−0.289, −0.098] | $7.4 \times 10^{-5}$ |
| pre-AD vs. post-AD | Entorhinal thickness | 194+73 | 0.516 | 0.752 | −0.237 [−0.325, −0.149] | $1.4 \times 10^{-7}$ |
| pre-AD vs. post-AD | Global conductance | 205+86 | 0.546 | 0.453 | +0.093 [−0.008, +0.194] | 0.070 |

On the headline pre-clinical contrast (CN vs. pre-AD), RCI significantly exceeds entorhinal thickness, global conductance, and a conventional DTI-ALPS index, and is statistically comparable to hippocampal volume (ΔAUC = +0.014, p = 0.49). In the symptom-onset analysis, once disease is clinically established, this advantage disappears: RCI does not significantly exceed the morphometric markers in the MCI track, and in the AD track it is significantly lower than both hippocampal volume (ΔAUC = −0.193) and entorhinal thickness (−0.237). RCI exceeds global conductance on the pre-clinical and MCI contrasts, and exceeds DTI-ALPS on the pre-clinical contrast (the task for which that comparator was included). The routing signal is thus specific to the pre-clinical window rather than to staging established disease.

## ADNI External Validation: Full Statistical Tables

ADNI primary analyses use the ComBat-harmonized full cohort ^40, 41^ (batch variable = acquisition protocol, preserving diagnosis/age/sex as biological covariates). Of 686 imaged sessions, 372 CN, 209 MCI, and 32 AD carried a diagnostic label; the harmonized conductance and RCI metrics are defined for 368 of the 372 CN (four CN dropped for missing conductance), giving the analysed cohort of n = 609 (368 CN, 209 MCI, 32 AD) reported in the harmonized Tier A rows below and plotted in Figure 7. As a scanner-confound sensitivity analysis we repeat the contrasts in the paired 2mm acquisition-protocol stratum (n = 412: 247 CN, 144 MCI, 21 AD), the largest homogeneous stratum, which eliminates scanner confound without harmonization; global conductance is defined for 245 of the 247 CN there (two CN lacked a computable conductance), so conductance contrasts use 245 CN and RCI contrasts use 247.

### Tier A: replication AUCs (DeLong 95% CI ^38^)

**Table S3:** ADNI Tier A replication: global conductance and RCI AUCs by diagnostic contrast and acquisition stratum. Global conductance is untrained, so its ADNI values are direct external validation. For RCI, the “frozen” rows apply the OASIS-trained weights unchanged to ADNI (true external validation); the remaining RCI rows re-estimate weights within ADNI and index reproducibility of the method rather than transfer of the trained model.

| Metric | Contrast | Stratum | AUC [95% CI] | $p$ |
| --- | --- | --- | --- | --- |
| Conductance | CN vs. MCI | paired_2mm | 0.667 [0.609, 0.725] | $4.0 \times 10^{-8}$ |
| Conductance | CN vs. MCI | full (ComBat) | 0.658 [0.611, 0.705] | $2.7 \times 10^{-10}$ |
| Conductance | CN vs. AD | paired_2mm | 0.815 [0.694, 0.935] | $1.7 \times 10^{-6}$ |
| Conductance | CN vs. AD | full (ComBat) | 0.805 [0.710, 0.900] | $1.0 \times 10^{-8}$ |
| Conductance | MCI vs. AD | paired_2mm | 0.678 [0.550, 0.806] | $8.5 \times 10^{-3}$ |
| Conductance | MCI vs. AD | full (ComBat) | 0.685 [0.581, 0.789] | $7.6 \times 10^{-4}$ |
| RCI | CN vs. MCI | paired_2mm | 0.817 [0.777, 0.858] | $1.2 \times 10^{-25}$ |
| RCI | CN vs. MCI | full (ComBat) | 0.830 [0.798, 0.862] | $9.5 \times 10^{-40}$ |
| RCI | CN vs. AD | paired_2mm | 0.898 [0.843, 0.953] | $1.4 \times 10^{-9}$ |
| RCI | CN vs. AD | full (ComBat) | 0.909 [0.867, 0.950] | $1.7 \times 10^{-14}$ |
| RCI | MCI vs. AD | paired_2mm | 0.633 [0.515, 0.750] | $5.0 \times 10^{-2}$ |
| RCI | MCI vs. AD | full (ComBat) | 0.642 [0.544, 0.740] | $9.7 \times 10^{-3}$ |
| RCI (frozen) | CN vs. MCI | OASIS→ADNI | 0.633 [0.585, 0.681] | $< 10^{-6}$ |
| RCI (frozen) | CN vs. AD | OASIS→ADNI | 0.797 [0.725, 0.868] | $< 10^{-12}$ |

**Table S4:** ADNI Tier B biological-specificity correlations. Positive correlation with CSF amyloid-*β*_42_ and negative correlation with pTau-217/amyloid-tau PET are the directions predicted under the glymphatic clearance hypothesis; the AQP4 and APOE4 dose-response rows are a specificity argument against a trivial genotype confound (see the main-text ADNI replication and Discussion). AQP4 dose is the allele count at rs12454744, immediately upstream of the AQP4 transcription start site (GRCh37 chr18:24,429,702, 2.3 kb 5*^′^* of AQP4) and within an astrocyte-specific super-enhancer in cell-type-resolved brain epigenomic maps^15^; *p* values are uncorrected for the small number of AQP4-region variants examined. Of the 28 correlations tested, the four full-cohort amyloid-PET and tau-PET rows are not shown.

| Predictor | Metric | Stratum | $\rho$ | $p$ ($n$) |
| --- | --- | --- | --- | --- |
| AQP4 rs12454744 dose | Conductance | paired_2mm | -0.017 | 0.841 (142) |
| AQP4 rs12454744 dose | RCI | paired_2mm | **0.196** | **0.020** (142) |
| AQP4 rs12454744 dose | Conductance | full cohort | 0.053 | 0.474 (186) |
| AQP4 rs12454744 dose | RCI | full cohort | **0.194** | **0.008** (186) |
| Amyloid PET SUVR | Conductance | paired_2mm | 0.162 | 0.241 (54) |
| Amyloid PET SUVR | RCI | paired_2mm | 0.009 | 0.947 (54) |
| Tau PET SUVR | Conductance | paired_2mm | -0.127 | 0.461 (36) |
| Tau PET SUVR | RCI | paired_2mm | -0.245 | 0.150 (36) |
| Plasma pTau-217 | Conductance | paired_2mm | -0.184 | 0.101 (81) |
| Plasma pTau-217 | RCI | paired_2mm | -0.235 | **0.035** (81) |
| Plasma pTau-217 | Conductance | full cohort | -0.189 | **0.030** (132) |
| Plasma pTau-217 | RCI | full cohort | -0.156 | 0.074 (132) |
| CSF amyloid-$\beta_{42}$ | Conductance | paired_2mm | **0.542** | **0.001** (34) |
| CSF amyloid-$\beta_{42}$ | RCI | paired_2mm | **0.499** | **0.003** (34) |
| CSF amyloid-$\beta_{42}$ | Conductance | full cohort | **0.484** | $1.4 \times 10^{-4}$ (57) |
| CSF amyloid-$\beta_{42}$ | RCI | full cohort | **0.397** | **0.002** (57) |
| CSF pTau | Conductance | paired_2mm | -0.015 | 0.931 (34) |
| CSF pTau | RCI | paired_2mm | 0.113 | 0.524 (34) |
| CSF pTau | Conductance | full cohort | 0.205 | 0.126 (57) |
| CSF pTau | RCI | full cohort | **0.365** | **0.005** (57) |
| APOE4 copy number | Conductance | paired_2mm | 0.070 | 0.238 (284) |
| APOE4 copy number | RCI | paired_2mm | 0.013 | 0.829 (286) |
| APOE4 copy number | Conductance | full cohort | 0.059 | 0.234 (414) |
| APOE4 copy number | RCI | full cohort | 0.044 | 0.372 (418) |

### Tier B: biological specificity (Spearman partial correlations, age/sex-adjusted) PET/plasma-to-DTI acquisition gap

**Table S5:** Interval between each ADNI biomarker’s acquisition date and the DTI session used in this study (nearest-date match, no ceiling on gap length), restricted to the same CN/MCI/AD analysis population used throughout Tier A/B (Figure 7). All three biomarkers are drawn from the allprojects table and matched to the closest-dated DTI session per participant, which is often from a different, earlier study phase.

| Biomarker | *N* matched | Median gap (days) | IQR (days) |
| --- | --- | --- | --- |
| Amyloid PET (SUVR) | 95 | 2,585 | 2,188–4,155 |
| Tau PET (SUVR) | 63 | 2,282 | 2,120–2,591 |
| Plasma pTau-217 | 136 | 2,352 | 2,052–3,849 |

Gaps of several years are common because ADNI’s PET and plasma sub-studies were not always acquired concurrently with the DTI session; the Tier-B correlations in the main-text ADNI replication should therefore be read as cross-sectional structural associations between white-matter routing and biomarker levels drawn from a nearby but non-concurrent study visit, not as a same-day, concurrent-validity test.

#### ADNI-specific limitations

DTI-ALPS is not computable in ADNI: ADNI diffusion data are reconstructed with generalized q-sampling (GQI), storing orientation distribution function peaks (index0/1/2) rather than the tensor field (dir0) that DTI-ALPS requires. The DTI-ALPS index and the raw per-edge conductance export are correspondingly unavailable in ADNI; the head-to-head DTI-ALPS comparison (main-text Results and Supplementary Table S2) is therefore OASIS-3-only, where the DTI-ALPS index (following ^11^) discriminated CN from pre-AD essentially at chance (AUC = 0.518 [0.475, 0.561]), far below RCI’s 0.819 on the same matched sessions (Supplementary Table S2). ADNI’s amyloid PET, tau PET, and plasma pTau-217 measurements were drawn from prior study phases; the median interval between biomarker acquisition and the DTI session used here is reported in Supplementary Table S5 and should be treated as a structural association with prior-phase biomarkers rather than a concurrent-validity test.

## Machine Learning: Cross-Validation, Hyperparameters, and Leakage Control

### Feature matrix

The full feature set comprises 2,306 columns per session, assembled as follows: 316 normative z-scores each for seven edge-level features: effective conductance, radial diffusivity (RD), fractional anisotropy (FA), streamline count (log-scale), the per-edge RCI log-likelihood-ratio, the top-1 Yen-path score, and route entropy (7 × 316 = 2,212); 80 normative z-scores for mean traversal time (one per parcel); six demographic/global covariates (age, sex, APOE4 dose, APOE2 dose, global FA, and years of education); three FreeSurfer structural indices (intracranial-volume-normalized hippocampal volume, entorhinal cortical thickness, white-matter hyperintensity volume); the count of structural routes present; and four cognitive measures (Mini-Mental State Examination (MMSE) ^44^ and Clinical Dementia Rating Sum-of-Boxes (CDR-SB) ^45^ slopes and most-recent values, restricted to assessments at or before the MRI date). These sum to 2,212 + 80 + 6 + 3 + 1 + 4 = 2,306. A whole-brain scalar RCI (the mean of the per-edge RCI over all 316 routes) is reported throughout the main text as the headline biomarker, but enters this classifier matrix only through its 316 per-edge constituents.

### Cross-validation scheme

All models are evaluated under 25-fold repeated stratified GroupKFold cross-validation (5 splits × 5 repeats), grouped by subject so that sessions from the same individual never appear in both training and test folds, stratified by the binary conversion label. Synthetic Minority Over-sampling (SMOTE) ^36^ is applied within each training fold to the minority (converter) class, after the normative z-score model has been fitted on training-fold CN sessions, to avoid imputing the test distribution. Converters within 730 days of diagnosis are up-sampled at a reduced ratio (0.40) to avoid generating unrealistic synthetic samples close to diagnosis. RCI LLR weights are re-estimated from training-fold sessions in every fold, eliminating circularity between LLR estimation and model evaluation.

### Binary classifier

The primary binary model is a histogram-based gradient-boosting classifier – a standard, off-the-shelf algorithm (scikit-learn HistGradientBoostingClassifier ^35^), not a novel contribution of this work; the novel contribution is the structural-routing feature set it is trained on. Hyperparameters: maximum depth 5, learning rate 0.05, minimum samples per leaf 20, maximum iterations 500, trained on the full 2,306-feature matrix to predict any pre-clinical conversion (pre-MCI or pre-AD vs. stable CN). Out-of-fold predicted probabilities are pooled across all 25 folds for evaluation. After the RCI leakage correction, cross-validated ROC-AUC is 0.814 and PR-AUC is 0.696 (Table 1, “All pre-clinical” row); models omitting the cognitive covariates (but retaining FreeSurfer and demographic features; run before the RCI leakage correction) achieve ROC-AUC 0.802–0.823 across two cross-validation runs, indicating that predictive information is not carried by the cognitive measures alone.

### Ordinal Frank–Hall model

To capture disease stage rather than binary conversion, an ordinal classifier uses the Frank–Hall decomposition ^13^. Four trajectory classes are defined: class 0 (stable CN, *n* = 991), class 1 (pre-MCI, n = 230), class 2 (pre-AD, *n* = 205), and class 3 (confirmed disease: post-MCI + post-AD, *n* = 230). The decomposition trains *K* − 1 = 3 independent binary threshold classifiers, *f_k_* : *P*(*Y* ≥ *k*) for *k* = 1, 2, 3, and recovers class probabilities as

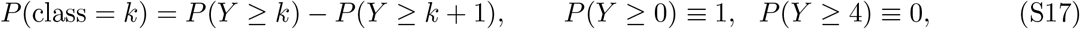

with predicted class *ŷ* = arg max*_k_ P*(class = k). Each threshold classifier is an ensemble of 10 HistGBT models trained on bootstrap resamples (30 models total). The model uses 24 curated features identified by SHAP importance ^37^ from the binary model (7 RCI edges, 3 entropy edges, 4 FA edges, 3 RD edges, 2 streamline-count edges, 2 mean-traversal-time regions, bilateral hippocampal volume and entorhinal thickness, and years of education). Age, sex, and APOE4 genotype are excluded: a feature-ablation analysis showed that adding them to the structural-routing features did not improve discrimination (ROC-AUC 0.812 with vs 0.813 without), indicating their demographic information is already absorbed by the structural-routing features. Pre-clinical ROC-AUC is 0.809, Spearman ρ = 0.536 (p = 1.0 × 10*^−^*^26^) between predicted ordinal score and true class, and 85.3% adjacent-class accuracy.

### Statistical reporting conventions

Group comparisons use Kruskal–Wallis tests for global effects and Mann–Whitney U with Bonferroni correction for pairwise comparisons. Ordinal trend across the five stages is quantified by the Kruskal–Wallis effect size ε^2^ = H/(n − 1), where H is the Kruskal–Wallis statistic and n the total sample size, and by the standardized Jonckheere–Terpstra statistic 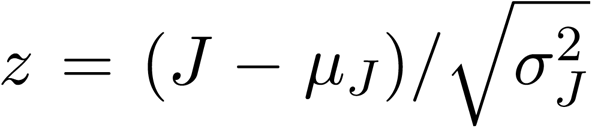 with 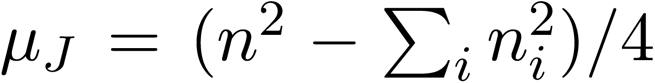 and 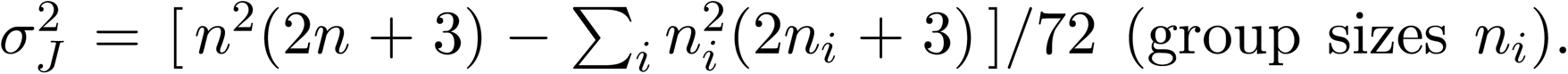 Effect sizes for two-group contrasts are reported as rank-biserial correlation *r*. Cohen’s d is not reported because Levene’s test indicated significant heteroskedasticity in most features. Two residualization procedures are used and should not be conflated: the head-to-head biomarker AUCs (Fig. 3, Supplementary Table S2) use an ordinary-least-squares residualization on age, sex, and APOE4 fitted on cognitively-normal sessions, whereas the cross-validated machine-learning classifier uses the fold-wise normative z-scoring of Eq. S15, which additionally log-transforms the edge features and adjusts conductance, radial diffusivity, and streamline count (but not RCI) for global FA. Classifier performance is reported as ROC-AUC and PR-AUC from pooled out-of-fold predictions, with DeLong 95% CIs ^38^. All analyses were conducted in Python 3.10 with scikit-learn 1.4 ^35^, scipy 1.12, numpy 1.26, and h5py 3.10.

## References

1 Reisa A. Sperling et al. “Toward Defining the Preclinical Stages of Alzheimer’s Disease: Recommendations from the National Institute on Aging-Alzheimer’s Association Workgroups on Diagnostic Guidelines for Alzheimer’s Disease”. In: Alzheimer’s & Dementia 7.3 (2011), pp. 280–292. doi: 10.1016/j.jalz.2011.03.003.

2 Clifford R. Jack et al. “NIA-AA Research Framework: Toward a Biological Definition of Alzheimer’s Disease”. In: Alzheimer’s & Dementia 14.4 (2018), pp. 535–562. doi: 10.1016/j.jalz.2018.02.018.

3 Andrew R. Hoy et al. “Microstructural white matter alterations in preclinical Alzheimer’s disease detected using free water elimination diffusion tensor imaging”. In: PLoS One 12.3 (2017), e0173982. doi: 10.1371/journal.pone.0173982.

4 Z. Alasmar, C. Tremblay, R. Moqadam, G. E. Serrano, T. G. Beach, A. Atri, Y. Su, Y. Zeighami, and M. Dadar. “Gray matter microstructure from in vivo diffusion magnetic resonance imaging reflects post mortem neuropathology severity and clinical progression of Alzheimer’s disease”. In: Alzheimer’s & Dementia 22.1 (2026), e71037. doi: 10.1002/alz.71037.

5 Andrea Coravos, Sean Khozin, and Kenneth D. Mandl. “Developing and Adopting Safe and Effective Digital Biomarkers to Improve Patient Outcomes”. In: npj Digital Medicine 2 (2019), p. 14. doi: 10.1038/s41746-019-0090-4.

6 Joanna M. Wardlaw et al. “Perivascular Spaces in the Brain: Anatomy, Physiology and Pathology”. In: Nature Reviews Neurology 16.3 (2020), pp. 137–153. doi: 10.1038/s41582-020-0312-z.

7 Jenna M. Tarasoff-Conway et al. “Clearance Systems in the Brain—Implications for Alzheimer Disease”. In: Nature Reviews Neurology 11.8 (2015), pp. 457–470. doi: 10.1038/nrneurol.2015.119.

8 Jeffrey J. Iliff et al. “A Paravascular Pathway Facilitates CSF Flow Through the Brain Parenchyma and the Clearance of Interstitial Solutes, Including Amyloid β”. In: Science Translational Medicine 4.147 (2012), 147ra111. doi: 10.1126/scitranslmed.3003748.

9 Maiken Nedergaard. “Garbage Truck of the Brain”. In: Science 340.6140 (2013), pp. 1529– 1530. doi: 10.1126/science.1240514.

10 Yuichi Chayama et al. “Physiological brain clearance architecture revealed by neuronal protein tracing”. In: Cell 189.15 (2026), pp. 4737–4755. doi: 10.1016/j.cell.2026.04.048.

11 Toshiaki Taoka, Yoshitaka Masutani, Hisashi Kawai, Toshiki Nakane, Kiwamu Matsuoka, Fumihiko Yasuno, Toshifumi Kishimoto, and Shinji Naganawa. “Evaluation of Glymphatic System Activity with the Diffusion MR Technique: Diffusion Tensor Image Analysis Along the Perivascular Space (DTI-ALPS) in Alzheimer’s Disease Cases”. In: Japanese Journal of Radiology 35.4 (2017), pp. 172–178. doi: 10.1007/s11604-017-0617-z.

12 Heiko Braak and Eva Braak. “Neuropathological Stageing of Alzheimer-related Changes”. In: Acta Neuropathologica 82.4 (1991), pp. 239–259. doi: 10.1007/bf00308809.

13 Eibe Frank and Mark Hall. “A Simple Approach to Ordinal Classification”. In: Machine Learning: ECML 2001. Vol. 2167. Lecture Notes in Computer Science. 2001, pp. 145–156. doi: 10.1007/3-540-44795-4_13.

14 William E. Klunk et al. “The Centiloid Project: Standardizing Quantitative Amyloid Plaque Estimation by PET”. In: Alzheimer’s & Dementia 11.1 (2015), pp. 1–15. doi: 10.1016/j.jalz.2014.07.003.

15 Alexi Nott et al. “Brain Cell Type–Specific Enhancer–Promoter Interactome Maps and Disease-Risk Association”. In: Science 366.6469 (2019), pp. 1134–1139. doi: 10.1126/science.aay0793.

16 Sibel Tekin and Jeffrey L. Cummings. “Frontal–subcortical Neuronal Circuits and Clinical Neuropsychiatry: An Update”. In: Journal of Psychosomatic Research 53.2 (2002), pp. 647– 654. doi: 10.1016/S0022-3999(02)00428-2.

17 Raphael M. Bonelli and Jeffrey L. Cummings. “Frontal-subcortical Circuitry and Behavior”. In: Dialogues in Clinical Neuroscience 9.2 (2007), pp. 141–151. doi: 10.31887/DCNS.2007.9.2/rbonelli.

18 Erlend A. Nagelhus and Ole P. Ottersen. “Physiological Roles of Aquaporin-4 in Brain”. In: Physiological Reviews 93.4 (2013), pp. 1543–1562. doi: 10.1152/physrev.00011.2013.

19 U.S. Food and Drug Administration. Enrichment Strategies for Clinical Trials to Support Determination of Effectiveness of Human Drugs and Biological Products: Guidance for Industry. U.S. Department of Health and Human Services. 2019. url: https://www.fda.gov/media/121320/download.

20 Pamela J. LaMontagne, et al. “OASIS-3: Longitudinal Neuroimaging, Clinical, and Cognitive Dataset for Normal Aging and Alzheimer Disease”. In: medRxiv (2019). doi: 10.1101/2019.12.13.19014902.

21 Ronald C. Petersen et al. “Alzheimer’s Disease Neuroimaging Initiative (ADNI): Clinical Characterization”. In: Neurology 74.3 (2010), pp. 201–209. doi: 10.1212/wnl.0b013e3181cb3e25.

22 Clifford R. Jack et al. “The Alzheimer’s Disease Neuroimaging Initiative (ADNI): MRI Methods”. In: Journal of Magnetic Resonance Imaging 27.4 (2008), pp. 685–691. doi: 10.1002/jmri.21049.

23 Peter J. Basser, James Mattiello, and Denis LeBihan. “MR Diffusion Tensor Spectroscopy and Imaging”. In: Biophysical Journal 66.1 (1994), pp. 259–267. doi: 10.1016/s0006-3495(94)80775-1.

24 Carlo Pierpaoli and Peter J. Basser. “Toward a Quantitative Assessment of Diffusion Anisotropy”. In: Magnetic Resonance in Medicine 36.6 (1996), pp. 893–906. doi: 10.1002/mrm.1910360612.

25 Rahul S. Desikan et al. “An Automated Labeling System for Subdividing the Human Cerebral Cortex on MRI Scans into Gyral Based Regions of Interest”. In: NeuroImage 31.3 (2006), pp. 968–980. doi: 10.1016/j.neuroimage.2006.01.021.

26 Arno Klein and Jason Tourville. “101 Labeled Brain Images and a Consistent Human Cortical Labeling Protocol”. In: Frontiers in Neuroscience 6 (2012), p. 171. doi: 10.3389/fnins.2012.00171.

27 Bruce Fischl et al. “Whole Brain Segmentation: Automated Labeling of Neuroanatomical Structures in the Human Brain”. In: Neuron 33.3 (2002), pp. 341–355. doi: 10.1016/s0896-6273(02)00569-x.

28 Fang-Cheng Yeh, Timothy D. Verstynen, Yibao Wang, Juan C. Fernández-Miranda, and Wen-Yih Isaac Tseng. “Deterministic Diffusion Fiber Tracking Improved by Quantitative Anisotropy”. In: PLoS ONE 8.11 (2013), e80713. doi: 10.1371/journal.pone.0080713.

29 Fang-Cheng Yeh, Van Jay Wedeen, and Wen-Yih Isaac Tseng. “Generalized Q-Sampling Imaging”. In: IEEE Transactions on Medical Imaging 29.9 (2010), pp. 1626–1635. doi: 10.1109/ TMI.2010.2045126.

30 Jin Y. Yen. “Finding the K Shortest Loopless Paths in a Network”. In: Management Science 17.11 (1971), pp. 712–716. doi: 10.1287/mnsc.17.11.712.

31 Edsger W. Dijkstra. “A Note on Two Problems in Connexion with Graphs”. In: Numerische Mathematik 1.1 (1959), pp. 269–271. doi: 10.1007/bf01386390.

32 Douglas J. Klein and Milan Randić. “Resistance Distance”. In: Journal of Mathematical Chemistry 12.1 (1993), pp. 81–95. doi: 10.1007/bf01164627.

33 Peter G. Doyle and J. Laurie Snell. *Random Walks and Electric Networks*. Vol. 22. Carus Mathematical Monographs. Mathematical Association of America, 1984.

34 Mikail Rubinov and Olaf Sporns. “Complex Network Measures of Brain Connectivity: Uses and Interpretations”. In: NeuroImage 52.3 (2010), pp. 1059–1069. doi: 10.1016/j.neuroimage.2009.10.003.

35 Fabian Pedregosa et al. “Scikit-learn: Machine Learning in Python”. In: Journal of Machine Learning Research 12 (2011), pp. 2825–2830. url: https://jmlr.org/papers/v12/pedregosa11a.html.

36 Nitesh V. Chawla, Kevin W. Bowyer, Lawrence O. Hall, and W. Philip Kegelmeyer. “SMOTE: Synthetic Minority Over-sampling Technique”. In: Journal of Artificial Intelligence Research 16 (2002), pp. 321–357. doi: 10.1613/jair.953.

37 Scott M. Lundberg and Su-In Lee. “A Unified Approach to Interpreting Model Predictions”. In: Advances in Neural Information Processing Systems. Vol. 30. 2017, pp. 4765–4774. url: https://proceedings.neurips.cc/paper/2017/hash/8a20a8621978632d76c43dfd28b67767-Abstract.html.

38 Elizabeth R. DeLong, David M. DeLong, and Daniel L. Clarke-Pearson. “Comparing the Areas Under Two or More Correlated Receiver Operating Characteristic Curves: A Nonparametric Approach”. In: Biometrics 44.3 (1988), pp. 837–845. doi: 10.2307/2531595.

39 Peter C. Austin. “Optimal Caliper Widths for Propensity-score Matching When Estimating Differences in Means and Differences in Proportions in Observational Studies”. In: Pharmaceutical Statistics 10.2 (2011), pp. 150–161. doi: 10.1002/pst.433.

40 W. Evan Johnson, Cheng Li, and Ariel Rabinovic. “Adjusting Batch Effects in Microarray Expression Data Using Empirical Bayes Methods”. In: Biostatistics 8.1 (2007), pp. 118–127. doi: 10.1093/biostatistics/kxj037.

41 Jean-Philippe Fortin et al. “Harmonization of Multi-site Diffusion Tensor Imaging Data”. In: NeuroImage 161 (2017), pp. 149–170. doi: 10.1016/j.neuroimage.2017.08.047.

42 Harold Jeffreys. “An Invariant Form for the Prior Probability in Estimation Problems”. In: Proceedings of the Royal Society of London. Series A, Mathematical and Physical Sciences 186.1007 (1946), pp. 453–461. doi: 10.1098/rspa.1946.0056.

43 Sheng-Kwei Song, Shu-Wei Sun, Michael J. Ramsbottom, Chen Chang, John Russell, and Anne H. Cross. “Dysmyelination Revealed Through MRI as Increased Radial (but Unchanged Axial) Diffusion of Water”. In: NeuroImage 17.3 (2002), pp. 1429–1436. doi: 10.1006/nimg.2002.1267.

44 Marshal F. Folstein, Susan E. Folstein, and Paul R. McHugh. ““Mini-mental State”: A Practical Method for Grading the Cognitive State of Patients for the Clinician”. In: Journal of Psychiatric Research 12.3 (1975), pp. 189–198. doi: 10.1016/0022-3956(75)90026-6.

45 John C. Morris. “The Clinical Dementia Rating (CDR): Current Version and Scoring Rules”. In: Neurology 43.11 (1993), pp. 2412–2414. doi: 10.1212/wnl.43.11.2412-a.

